# Longitudinal degeneration of microstructural and structural connectivity patterns following stroke

**DOI:** 10.1101/2025.05.11.25327333

**Authors:** Lorenzo Pini, Mohammad Hadi Aarabi, Alessandro Salvalaggio, Angela M. Colletta, Nicholas V Metcalf, Joseph C Griffis, Alex R Carter, Gordon L. Shulman, Maurizio Corbetta

## Abstract

Stroke leads to neurological impairment through local and widespread brain damage. Whether distal and local diffusivity changes follow similar reorganization and behavior correlation trajectories is unclear. We examined acute and chronic brain diffusivity changes in stroke patients using both connectivity and microstructural parameters, hypothesizing similar trajectories linked with behavioral scores. This perspective study assessed first-time stroke patients at two weeks and three months using behavioral tests and diffusion MRI. We applied latent factorial analysis to behavioral and microstructural data from diffusion tensor imaging (DTI) and neurite orientation dispersion and density imaging (NODDI) outcomes. Structural connectivity gradients were computed from whole brain tractography. Statistical analyses included cross-sectional analyses, longitudinal linear mixed models, and the assessment of the relationship between microstructure and contralateral dysconnectivity. Finally, we explored the linear relationships between diffusivity parameters and behavior. Seventy-nine patients (60±12 years) were enrolled, with 32 completing follow-up (3 months). A healthy group of n=33 (58±11 years) was included. Factorial analysis identified five latent factors explaining about 50% of behavioural variances. We described three main DTI-NODDI microstructural maps and three main structural gradients, capturing around 50% of variance each. The analysis revealed acute alterations of microstructural and connectivity patterns. Longitudinally, significant degeneration in structural connectivity was observed, echoed in microstructural parameters. Behaviourally, global structural connectivity alterations were associated with acute cognitive deficits, though this link weakened at chronic follow-up. On the contrary, local disconnectivity patterns were linked mainly with motor deficits. Finally, we reported a significant association between global connectivity alterations and microstructural changes in tracts locally disconnected. Stroke significantly alters diffusivity patterns beyond the lesion, with no recovery over time. While acute-stage effects relate to behavior, long-term connectivity changes seem mostly uncoupled, suggesting they do not correlate with stroke recovery.

## 1. INTRODUCTION

Stroke is the second leading cause of death (about 10-12% of all deaths), and the first cause of disability worldwide.^1,2^ After a stroke, a cascade of cellular events unfolds in both the lesion and perilesional areas. Within the lesion, compromised blood supply triggers neural damage, disrupting cellular homeostasis.^3^ Immediate responses include excitotoxicity, oxidative stress, and inflammation, leading to neuronal injury and death.^4–7^ In the perilesional area, while hypoperfusion and hypoxia can cause damage, these effects may be reversible.^8–11^ Animal studies have shown that the perilesional region undergoes dynamic changes in cellular composition, synaptic connectivity, and gliosis.^12^ Importantly, both functional and structural changes occur at distances from the lesion, and these remote changes have been associated with behavioral deficits or recovery.^13–16^

The post-stroke pattern brain alterations are complex due to the nature of the brain’s connectome, that is the macro-scale brain connectivity properties, both functional (post-synaptic) and structural (axonal) pathways. Several studies have documented changes in the connectome after a stroke. Our group has pioneered methods to study alterations in functional connectivity (FC) measured by resting-state functional MRI between brain regions,^17–20^ known as ‘connectional diaschisis’.^21^ Stroke lesions, regardless of location, cause characteristic patterns of FC alterations, including decreased inter-hemispheric correlation between homotopic regions and decreased intra-hemispheric negative correlation between typically segregated networks.^20,22^ These changes explain behavioral impairments across multiple domains and, when normalized over time, account for functional recovery.^15,23,24^ We also showed that functional changes are correlated with changes in structural connectivity (SC), albeit indirectly measured by embedding lesions in healthy subject atlases of white matter (WM) connections.^25,26^ Embedding lesions into normative atlases of functional/structural connections to explore the pattern of disconnection has become a powerful method to infer the effect of lesions onto brain networks.^27–29^

While FC changes are well-documented, much less is known about directly measured alterations of the structural connectome and microstructure after focal stroke injury and recovery.^30–33^ In humans, diffusion magnetic resonance imaging (dMRI) enables detection of structural alterations by sensitizing the MRI signal to water movement as a proxy of WM properties.^34–36^ Mathematical models like diffusion tensor imaging (DTI) and biological models like neurite orientation dispersion and density imaging (NODDI) provide detailed insights into tissue microstructure ^36–38^.

SC and microstructure studies in stroke have demonstrated both sub-acute and chronic changes in cross-sectional studies.^39–46^ Interestingly, while FC tends to normalize over time in relation to stroke recovery,^15,23,47,48^ longitudinal studies have shown progressive segregation of SC and WM degeneration across the brain post-stroke.^30,32,33^

However, the spatial and temporal evolution of these structural changes, the relationship with behavior, and, particularly, how alterations in SC patterns are related to microstructural changes, remain poorly understood.

The aim of this study was to measure the effect of stroke lesion on both SC and microstructural parameters. We focused on measures of whole-brain SC from diffusion imaging in the damaged and intact hemisphere, based on the inter- and intra-hemispheric functional changes reported in previous studies^20,49,15^, as well as on the microstructural diffusion parameters measured in the WM tracts and cortical gray matter disconnected by the lesion, based on a growing literature on the importance of disconnection in accounting for post-stroke behavioral deficits.^27,50–53^ Furthermore, we measured both diffusion MRI and behavior with an in-depth behavioral battery that explored multiple domains of function^23,54^ at two time points: sub acutely within 2 weeks from the stroke and chronically at 3 months when most recovery has occurred.^47,55,56^ We were interested in two main questions. First, given previous work showing a robust coupling between network-level functional MRI alterations and structural disconnection *indirectly* measured through healthy atlases,^26^ we were interested in *directly* measuring alterations of whole brain SC and diffusion microstructure in WM and gray matter disconnected by the lesion. While previous studies have focused on graph metrics of diffusion changes post-stroke,^30,32,33^ this is the first study to combine measures of SC and microstructure both at the whole brain level and in regions disconnected by the lesion. Second, given that animal models suggest remodeling of WM pathways in support of recovery of function,^57^ and human data indicate an important role of functional network normalization,^15,23,47,48^ we were interested in measuring changes of whole brain SC and microstructure of disconnected tissue longitudinally, and their relationships with behavioral recovery. A positive relationship would support a role of WM reorganization and/or remodeling in mediating recovery of function, while no relationship would suggest that behavioral recovery is relatively independent of structural remodeling.

## 2. METHODS

### Study design and participants

Patients with first-time stroke were enrolled at Washington University in Saint Louis and were prospectively followed there at two-week three-month, and one-year intervals from the stroke (data from the 1-year timepoint are not reported in the present study). Inclusion and exclusion criteria are listed in **Supplementary Material S1.1**. At each visit, patients underwent behavioral testing and MRI examination. Healthy controls (HC) were matched with the patients by age, sex, and years of education, and were studied twice one month apart. At the baseline and follow-up visits, patients and HC underwent the same behavioral testing and MRI scans. The workflow of the study is depicted in **Fig. 1**. Written informed consent was obtained from all participants in accordance with procedures established by the Washington University in Saint Louis Institutional Review Board.

**Figure 1.**
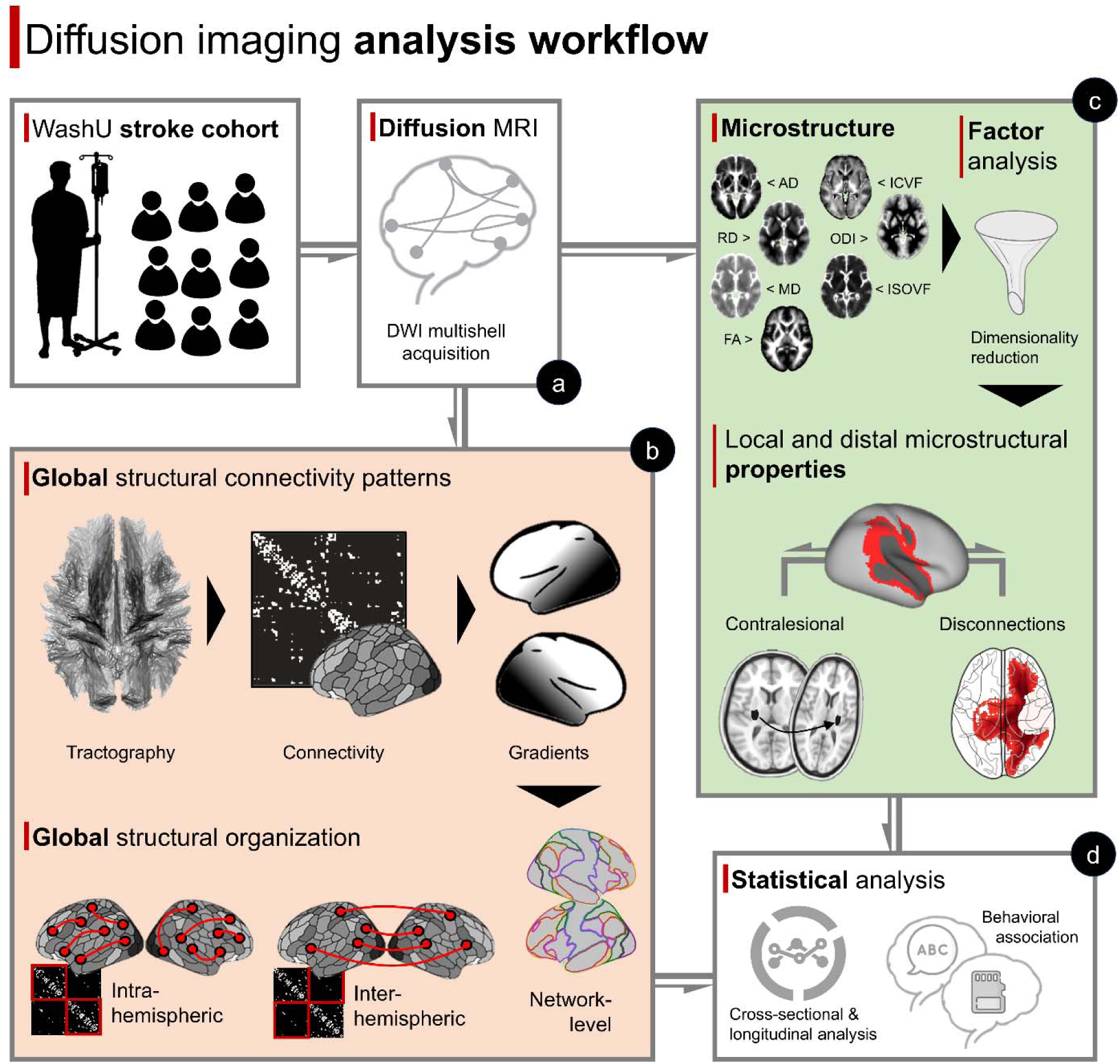
Overview of the analysis. **Lef panels. Panel A**: Stroke and age-matched controls underwent diffusion imaging multi-shell. Two different analyses were performed: **Panels B**: whole brain structural connectivity matrices were computed by tractography. Next, a diffusion embedding algorithm computed connectivity gradients separately for intra- and inter-hemispheric connections both at the parcel level and network level ^63^. **Panels C**: the same diffusion data were used to compute microstructural models (diffusion tensor imaging and neurite orientation dispersion and density imaging). The resulting maps were fed into a factorial analysis to measure microstructure of white matter and gray matter of disconnected tracts and contralesional regions (homotopic to the lesion). **Panels D**: Connectivity (gradients) and microstructural patterns were compared cross-sectionally (controls vs. stroke), longitudinally (2 weeks vs. 3-month follow-up), and for linear relationship with behavioral scores.

### Behavioral assessment

Participants underwent an extensive behavioral battery, administered typically on the same day as the imaging session and lasting approximately 2.5 hours, representing an update of a validated battery used in a previous cohort ^54^. The following cognitive domains were assessed: memory (Hopkins Verbal Learning Test-Revised; Brief Visuospatial Memory Test-Revised; WMS-III Spatial Span, forward and backward); language (Boston diagnostic aphasia examination; nonword reading, subtest of the Delis-Kaplan executive function system - animal naming), visuospatial attention functions (Posner spatial cueing task, Mesulam Symbol Cancellation Test), executive functions (sorting card test and flanker test from the NIH Toolbox), and motor abilities (Action Research Arm Test, combined walking Index, lower extremity Motricity Index). In line with our previous publications ^54,58,59^, a dimensional reduction approach was applied by means of a hierarchical factor analysis with orthogonal (varimax) rotation to explore the latent variable structure. The resulting cognitive factors were compared between stroke control groups using a two-sample t-test, and between the sub-acute and chronic stroke phases with a paired t-test. A full description of sub-scores and processing details are reported in **Supplementary Materials S1.2**.

### MRI acquisition and preprocessing

A 3 T Siemens Prisma scanner collected neuroimaging data. The anatomical sequence was 3D T1 Magnetization Prepared Rapid Acquisition Gradient Echo (MP2RAGE) (echo time/repetition time/inversion time= 236 /1700/1000 ms, 176 axial slices, 1-mm slices, 240-mm FOV, 256[×[256 acquisition matrix), 3D FLAIR (repetition time/echo time/inversion time = 5000/394/1800 ms, 1 mm3 isotropic spatial resolution) and 3D T2 (repetition time/echo time = 32/564 ms, 1 mm3 isotropic spatial resolution). A multi-shell diffusion scheme was acquired via a whole-brain coverage, and the b-values were 0, 300, 1000, and 2000 s/mm2. The diffusion sampling directions were 16, 16, 32, and 60, respectively. The in-plane resolution was 2 mm with a slice thickness of 2 mm. Resting-state and task functional MRI scans were also collected but are not included in the present study. Preprocessing of the diffusion data is reported in **Supplementary Materials S1.3.**

### Global SC and local microstructural patterns computation

#### Global patterns: connectivity gradients

The MICAPIPE pipeline ^60^ was applied to compute the SC matrix from T1w and dMRI preprocessed data. A connectivity matrix was used to generate cortex-wide structural connectome gradients by applying diffusion map embedding algorithm, a non-linear approach which preserves the local geometry of the data and is robust against noise ^61^. Details are reported in **Supplementary material S1.4**. In line with the literature ^62^, we *a priori* selected the first 3 principal gradients, to provide a balanced representation of connectivity pattern, sufficiently detailed to capture the complexity of the data.

To quantify the variability of the main gradients included in the analysis, we applied a gradient divergence (GD) measure for each brain parcel. First, we computed the group averaged- gradient maps from the normative sample; then for each patient and gradient we computed parcel-wise Euclidean distance, representing the distance between each parcel gradient value and the normative value from the healthy gradient templates, according to the following formula:

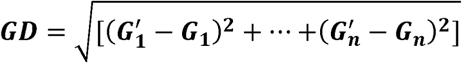

where n represents the number of gradients retained from the analysis and G’ the gradient value at the parcel level from the reference template. Finally, the GD values were normalized between 0 and 1 (with a min-max scaling approach). GD was computed separately for intra- and inter-hemispheric gradients. For the network-wise analysis GD was averaged across Yeo’s network template^63^ (default mode network: DMN; dorsal-attention network: DAN; ventral-attention network: VAN; frontoparietal network: FPN; visual network: VIS; sensorimotor network: SMN; limbic network: LIM).

Gradients values were computed also at network levels (average gradient score), according to the matching between Shaefer’s parcellation and 7 Yeo’s network template.

Finally, to visualize the main connectivity features of the structural gradients, we assessed the relationship between the first two main gradients identifying the extreme parcels (from the control group) in the cartesian axis (left, right, up, and bottom) within specific network. The structural pathways originating from these cartesian points for both intra- and inter-hemispheric connectivity were plotted from a normative streamline map ^64^.

#### Local-dysconnectivity patterns: microstructural space

Diffusion processed data were used to estimate both DTI and NODDI measurements. For DTI metrics, Dipy was employed ^65^. Diffusion data were truncated at b=1000 and the following DTI metrics were fitted: fractional anisotropy (FA), mean diffusivity (MD), axial diffusivity (AD), and radial diffusivity (RD). For NODDI, we employed the Accelerated Microstructure Imaging via Convex Optimization framework ^66^. We derived maps for neurite density index (NDI), orientation dispersion index (ODI), and isotropic volume fraction (ISOVF). Both DTI and NODDI maps were fed into a latent factor analysis. This analysis was performed at both individual-space and group-average level normalized to the MNI template (**Supplementary material S1.5**). Due to the exploratory nature of this latent DTI and NODDI approach, the same method was applied in an independent sample of n=1064 individuals from the Human Connectome Project (HCP) using multi-shell diffusion data (preprocessing details are reported in **Supplementary materials S1.6**).

Microstructural analyses were focused exclusively on i) contralesional GM regions, and ii) WM tracts structurally disconnected from the lesion. Specifically, the lesion mask, binarized and registered to the MNI was flipped in the y-axis to derive the contralesional region. ROIs were further divided into GM and WM using the Harvard-Oxford atlas (25% probability map) as a mask. Microstructural latent profiles were assessed also at tract levels, considering the structural disconnection (SDC) patterns. Specifically, for each patient the corresponding structural disconnection map ^50^ was used as a mask to derive distal-connected averaged factorial values. Structural disconnection maps within the stroke sample were described through a principal component analysis and the components explaining more than 10% of the variance plotted as tractography maps for visualization purposes. We compared three different disconnection probability thresholds (40, 60, and 80%) to assess the robustness of the analysis. To avoid partial volume effect in the structural disconnection maps, we also computed a “*shell*” region surrounding the lesion which was removed from the tracts: an exclusion region map was created inflating of 4 voxels the lesion borders.

### Cross-sectional comparison between sub-acute stroke and controls

#### Global connectivity differences

First, we assessed whether the distributions of the relationship between the main gradients were significantly different between cohorts. We applied structural equation modeling (SEM) to investigate the relationship between brain gradient metrics across different brain networks (details in **Supplementary materials S1.7**). For this analysis, we assessed whether group membership (HC vs Stroke) influenced the association between these two variables. Each network in the dataset was analyzed separately to determine network-specific patterns. Similarly, we intra- and inter-connectivity gradients were analyzed independently.

To summarize and condense the information from gradient values and brain parcels, data were projected into a two-dimensional (2D) space using the Uniform Manifold Approximation and Projection (UMAP), a non-linear embedding technique that efficiently represents high-dimensional data by preserving local and global structure along the principal axes ^67^. Specifically, for each individual (both stroke patients and controls), gradient values from each parcel were processed through UMAP, reducing the dimensionality of the data from *parcels × gradients* to two principal components per subject. UMAP default parameters were applied as in our prior work ^68^. Following this dimensionality reduction, a multivariate analysis of covariance (MANCOVA) was conducted to examine group differences (stroke vs. controls) in the 2D UMAP space. The MANCOVA model included group as the main factor, controlled for covariates (age, education, and gender) to account for potential demographic effects. This approach enabled the isolation of group effects while controlling covariate influence. Further, a support vector machine (SVM) was applied to assess the accuracy of classifying controls and stroke (sub-acute) in the UMAP space. To this aim, the dataset was split into training (65%) and test (35%) set for hyperparameter validation with a Leave-One-Out Cross-Validation strategy. The hyperparameter grid explored included variations in the regularization parameter C tested across a range of values from 0.1 to 10, incremented by 0.2 (C=[0.1,0.3,0.5,…,9.9]), kernel types and gamma values. The accuracy of the model was assessed in the test set.

Since the GD metric was derived from the control group data, a direct comparison between controls and stroke patients was not conducted. Instead, a one-sample t-test was performed across stroke patients to identify parcels where GD values mainly deviated from zero. Here, zero represents a perfect alignment with the control group’s GD template, so positive t-scores indicate areas where the stroke group’s GD values differ from this baseline. For visualization, only parcels with t-scores exceeding 5 were plotted, highlighting the regions with the most pronounced deviations from the control-derived GD.

Finally, we compared stroke and HC data from session 1 using directly gradient values. For stroke patients, the hemispheres ipsilesional to the lesion were flipped to the left hemisphere (when needed), while the right hemisphere was considered as the contralesional to the lesion hemisphere. Each parcel was classified as the corresponding functional Yeo’s 7 networks. An absolute value was computed for each parcel before computing the network average value (to prevent extreme and opposite values in parcels belonging to the same network from skewing the mean towards 0 in one of the two groups). We applied an ordinal least square regression (OLS) to compare stroke and HC (session 1) covarying for age, gender, and education at network level. We tested the effects of diagnosis and networks, as well as the interaction effect (*diagnosis × network*) using network average gradient scores for each of the seven Yeo’s networks ^63^.

#### Local disconnectivity differences

Latent microstructural diffusion factors within SDC maps were compared between stroke and controls. First, we created a mean template from the control group for the latent factor maps. For each patient, the corresponding SDC masks were used as ROI in the control’s template. Values within the individual and template maps were compared by means of non-parametric independent test (Mann–Whitney U test), with significance set at p<0.05. Further, we applied a voxel-wise analysis comparing latent factors between controls and stroke (sub-acute), limited to the WM mask (from the Harvard-Oxford atlas – 25% probability map), described in **Supplementary Materials S1.8**.

#### Disconnectivity probability and microstructural outcomes

Further, we assessed whether the contralesional microstructural latent parameters could predict the likelihood of the contralesional area being disconnected from the lesion. To this aim, a linear regression analysis was applied including as independent variables all the factorial values within the GM contralesional tissues (DTI and NODDI) and the structural degree of dysconnectivity as the dependent variable. A bootstrapping approach (n=1000) was applied for the regression analysis to assess the robustness of the prediction (R^2^) value. Moreover, results from the bootstrapping procedure were used to compute F-statistics linked with the R^2^ distribution within each model, to assess the stability of the regression within the stroke sample according to the following formula:

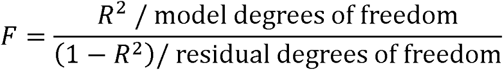

In a second analysis, we investigated whether GM regions disconnected from the lesions exhibited distinct diffusion values compared to unconnected regions at the sub-acute stage. Utilizing the SDC maps without applying a probability threshold, we calculated the probability of disconnections for each GM parcel (n=200 according to ^69^, including cerebellum parcels from Buckner atlas^70^. The factorial values averaged within the parcel exhibiting the highest probability were compared with those within the parcel displaying the lowest probability, employing a paired t-test. Given that multiple parcels can share the lowest probability (equal to 0), we randomly selected a parcel with the lowest probability and repeated the procedure n=100 times. Each time, we compared the randomly selected low-value parcel with the single parcel exhibiting the highest value. In each iteration, the corresponding statistics (p-value) were recorded, and for each diffusion factor identified by the latent DTI-NODDI model, we compared the respective values. This approach was repeated both including and excluding subcortical regions (from the Oxford template) to avoid potential biases linked with the distribution of the lesions.

### Longitudinal analysis: sub-acute vs chronic stroke

We employed a linear mixed model (LMM) analysis for comparing GD values longitudinally in stroke patients (sub-acute vs 3 months follow-up). The analysis was performed at network level (parcel-GD average), considering networks and individuals as random factors. This analysis was repeated for both intra- and inter-connectivity structural gradients and for ipsi- and contralesional hemispheres (for a total of n=4 models). For assessing the robustness of results, the analysis was performed both on the whole dataset (intention-to-treat like; ITT-L) and on the subset of patients with available dMRI data for both sub-acute and follow-up (per-protocol-analysis like; PPA-L). Finally, we compared microstructural latent factors longitudinally (sub-acute vs chronic) within the lesion and SDC masks, using the same LMM applied for gradients. As for the above analysis, we applied the model for both ITT-L and PPA-L samples. The main sociodemographic factors were covaried from the analysis for both GD and latent microstructural outcomes.

### Global and local connectivity association with behavior

We assessed the relationships between behavioral latent factors with intra- and inter-hemispheric GD for sub-acute and chronic stages, separately. Specifically, for each cognitive factor, we assessed the linear relationship (Pearson’s correlation) with each structural gradient parcel-wise. This analysis was performed independently for intra- and inter-connectivity and significance was set at p<0.05 FDR corrected.

The relationship between microstructural properties and behavioral performance was assessed by means of a linear regression approach. Specifically, latent diffusion values within SDC and lesion masks were included as predictors for the cognitive latent factors. The analysis was repeated independently for sub-acute and chronic stages. A p-value Bonferroni corrected for the number of behavioral scores was considered significant. Outliers exceeding two times the interquartile ranges were removed from the linear regression analysis to avoid the inclusion of potential extreme values that could bias the model. For SDC we considered only the more lenient threshold (40%).

### Global connectivity organization and microstructural parameters association

Finally, we assessed the relationship between gradient organization and microstructural parameters. Specifically, we aimed to relate whether changes from controls in the SC hierarchy (GD) were related to biological mechanisms captured by the microstructural space focusing on disconnected tracts. To this aim, we explored the relationship between global connectivity patterns (from structural gradients) with microstructural parameters in brain regions belonging to disconnected tracts (i.e., SDC) from the lesion. The DTI-NODDI factors computed within the SDC tracts were linearly related to the GD value, independent for intra and inter-connectivity and for the ipsilesional and contralesional hemisphere. First, we computed a global GD value averaging the value from each parcel. Microstructural parameters in the SDC ROI were used as predictors of the global GD. A p-value<0.05 was considered significant. Further, we perform this analysis at network level, averaging GD within the 7 Yeo’s network parcels. A p-value<0.05 FDR-corrected was considered significant. This analysis was performed in sub-acute stroke patients. This analysis was performed considering SDC maps thresholded at the lenient cut-off (40%).

### Sensitivity and reproducibility analyses

We performed several sensitivity analyses to check the robustness of cross-sectional, longitudinal and connectivity analyses. Further, we considered for the connectivity analysis the sample of patients with lesions involving mainly subcortical and non-subcortical regions. Details for these analyses are in **Supplementary materials S1.9**.

## 3. RESULTS

A total of 79 patients (age=60.1±11.5 years; education=13.3±2.5 years) and n=33 age-matched HC were included (**Table 1**). Patients and controls were matched for age (T=0.927, p=0.356), education (T=−0.897, p=0.372), and gender (X^2^=1.433, p=0.231). Among HC, n=29 individuals performed MRI exam (age=57±11; education=13.7±2.6), of whom n=22 repeated the MRI visit 1 month apart. A total of 48 patients had available MRI data at the baseline visit (age=59±11; education=13.5±2.2) while n=26 repeated the exam at the 3 months follow-up (age=59±11 years, education=13.8±2.2 years). As for the whole cohort, no differences were observed between the stroke baseline imaging cohort and the cohort of controls performing MRI at the first session (age: T=−0.55; p=0.58; education: T=0.38; p=0.71; gender: X^2^=3.06: p=0.08). An overview of the maps derived from diffusion-weighted imaging is presented in **Fig. 2**, alongside the lesion frequency map and a visual representation of the main SDC patterns obtained through PCA.

**Figure 2.**
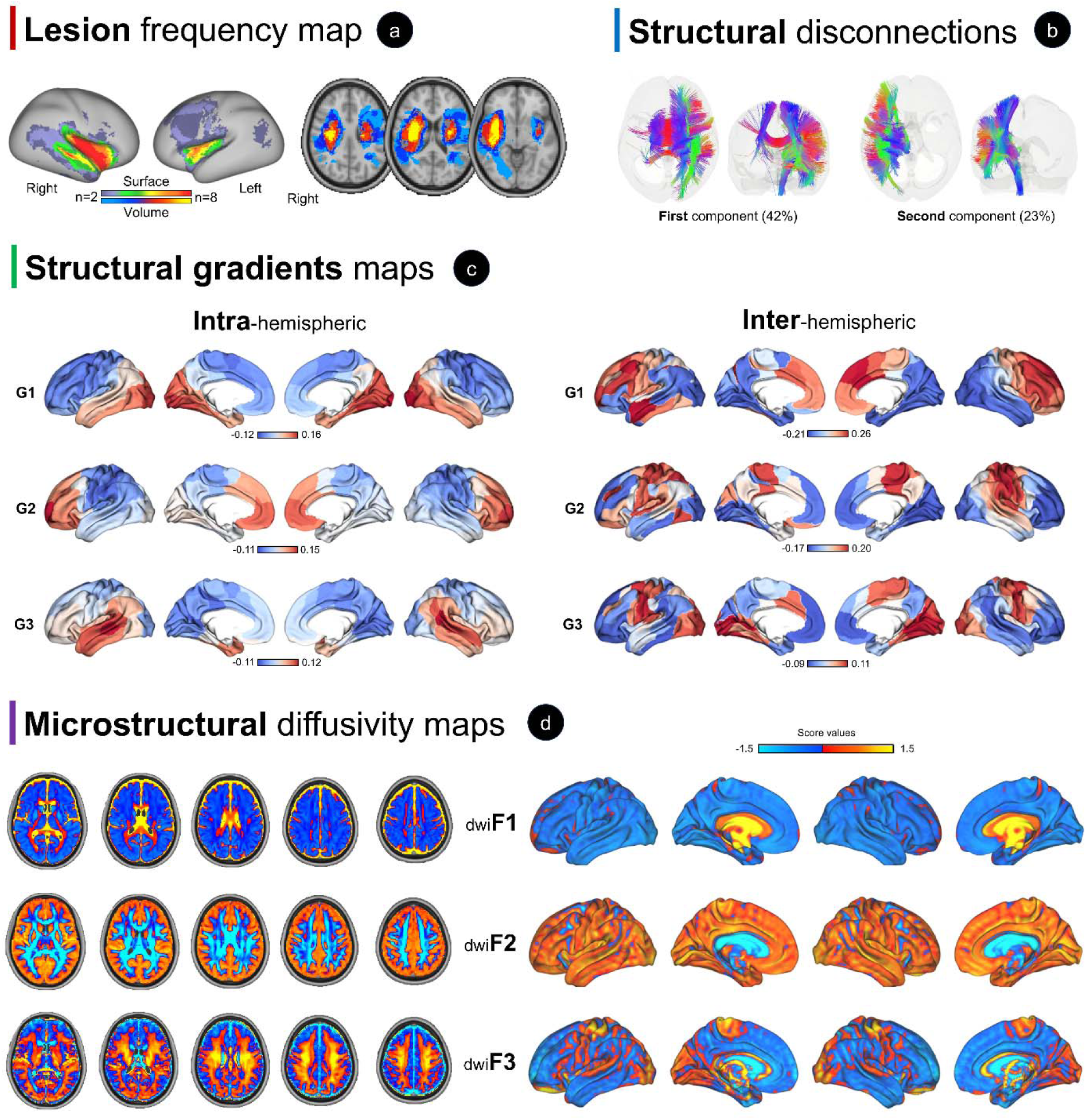
Brain imaging maps employed in the study. **Panel a**: lesion frequency map projected both at the surface and volume level. **Panel b**: structural disconnection patterns (the first two main components are shown at fiber-level and 3D rendering). All analyses were carried out on individually computed disconnection maps. **Panel c**: intra-hemispheric and (left panels) inter-hemispheric (right panels) connectivity gradients from the healthy control population (WashU cohort). **Panel d**: microstructural latent diffusion maps at both volumetric (left panels) and surface (right panels) space (healthy controls of the WashU cohort).

**Table 1.** Whole-cohort of stroke patients and healthy controls (intention-to-treat sample)

|  | Healthy controls | Stroke patients | p-values |
| --- | --- | --- | --- |
| **Sociodemographic** |  |  |  |
| Numerosity (ITT-L at T0) | 33 | 79 | / |
| Age | 57.9 ± 10.9 | 60.1 ± 11.5 | 0.36 |
| Education | 13.8 ± 2.5 | 13.3 ± 2.5 | 0.37 |
| Gender (Female) | 19 (58%) | 34 (43%) | 0.23 |
| **Behavioral assessment** |  |  |  |
| Flanker test | 104 ± 20 | 80 ± 21 | <0.001 |
| Card sorting test | 116 ± 24 | 91 ± 25 | <0.001 |
| Mesulam total missing |  |  |  |
| *total missing* | 1.69 ± 2.84 | 6.84 ± 14.5 | 0.05 |
| *center of cancellation* | -0.0015 ± 0.01 | 0.005 ± 0.21 | 0.85 |
| Posner test |  |  |  |
| *accuracy average* | 0.99 ± 0.02 | 0.86 ± 0.19 | <0.001 |
| *accuracy visual field* | -0.001 ± 0.04 | 0.092 ± 0.3 | 0.08 |
| *accuracy validity* | 0.006 ± 0.02 | -0.029 ± 0.09 | 0.02 |
| *accuracy disengage* | 0.003 ± 0.03 | 0.01 ± 0.09 | 0.64 |
| *reaction time average* | 527 ± 109 | 743 ± 358 | 0.001 |
| *reaction time visual field* | 2.47 ± 93 | 44 ± 488 | 0.63 |
| *reaction time validity* | -8.71 ± 56 | 5.08 ± 204 | 0.70 |
| *reaction time disengage* | 8.76 ± 50 | 21.9 ± 195 | 0.70 |
| Fluency test (animal) | 21.3 ± 4.78 | 16.9 ± 8.77 | 0.007 |
| Comprehension oral sentence | 36.6 ± 0.68 | 35.2 ± 4.2<sup>A</sup> | 0.079 |
| Reading oral sentences | 17.9 ± 3.2 | 14.9 ± 6.45 | 0.02 |
| Non word | 17.88 ± 3.21 | 14.92 ± 6.45 | 0.017 |
| Boston score | 13.6 ± 1.88 | 12.3 ± 3.63 | 0.07 |
| Brief Visuospatial Memory Test |  |  |  |
| *Percent retained* | 87.4 ± 22.8 | 74.5 ± 31.5 | 0.04 |
| *Total Immediate Recall* | 42.5 ± 12.9 | 28 ± 18.9 | <0.001 |
| *Discrimination Index* | 5.54 ± 0.90 | 4.55 ± 1.69 | 0.002 |
| *Delayed recall* | 45.2 ± 14.8 | 29.2 ± 19.9 | <0.001 |
| Hopkins Verbal Learning Test |  |  |  |
| *Percent retained* | 75.8 ± 24.6 | 56.5 ± 35.7 | 0.006 |
| *Total Immediate Recall* | 43.4 ± 13.2 | 28.8 ± 16.2 | <0.001 |
| *Discrimination Index* | 10.1 ± 1.45 | 7.92 ± 2.96 | <0.001 |
| *Delayed recall* | 38.2 ± 18.2 | 24.5 ± 20.2 | 0.001 |
| Digit span forward | 7.79 ± 2.26 | 6.25 ± 2.33 | 0.003 |
| Digit span Backward | 6.48 ± 2.00 | 5.05 ± 2.57 | 0.007 |
| Walk test |  |  |  |
| *Time* | 1.21 ± 0.42 | 0.52 ± 0.61 | <0.001 |
| *Functional independence* | 7 ± 0 | 4.96 ± 2.20 | <0.001 |
| *Combined walk test* | 8.88 ± 0.33 | 6.03 ± 2.95 | <0.001 |
| Motricity index |  |  |  |
| *Left lower extremity* | 99 ± 3.32 | 86.9 ± 23.5 | 0.004 |
| *Right lower extremity* | 92.3 ± 3.07 | 86.4 ± 24.8 | 0.004 |
| Action Research Arm |  |  |  |
| *Left grasp* | $18 \pm 0$ | $15.3 \pm 6.18$ | *0.014* |
| *Right grasp* | $18 \pm 0$ | $15.2 \pm 5.81$ | *0.007* |
| *Left grip* | $12 \pm 0$ | $10.1 \pm 4.17$ | *0.010* |
| *Right grip* | $12 \pm 0$ | $10.2 \pm 3.87$ | *0.008* |
| *Left pinch* | $18 \pm 0$ | $14.4 \pm 6.71$ | *0.003* |
| *Right pinch* | $18 \pm 0.17$ | $14.3 \pm 6.24$ | *0.001* |
| *Left gross* | $9 \pm 0$ | $8.01 \pm 2.74$ | *0.042* |
| *Right gross* | $9 \pm 0$ | $7.75 \pm 2.99$ | *0.019* |

A: missing on more than 25% of the stroke sample

### Latent behavioral factors

A dimensional reduction approach was applied to characterize the latent space of behavioral scores in both stroke and HC. The latent factorial analysis (Kaiser-Meyer-Olkin=0.813), after excluding individuals with missing data exceeding the established threshold (n=59 for patients in the sub-acute stage and n=35 for follow-up) revealed 5 main latent factors explaining around 50% of the behavioral variance. The first factor mainly loaded on verbal memory, language, and executive functions tests/items (variance=13.3%). The second (variance=12.2%) and third (variance=8.1%) factors loaded on right and left motor functions, respectively. The fourth (variance=6.4%) factor mapped onto visual memory, working memory, and general attention performance, while the fifth (variance=6.38%) related mainly to lateralized visuospatial scores on the Posner task. Due to the left-right nature of the last factor, we considered the absolute values. Scores from these factors were significantly different between HC and stroke at the sub-acute stage (p<0.05)(**Fig. 3**). The factorial analysis was repeated considering only the stroke (acute) sample, mirroring the results in the whole cohort (**Supplementary Fig. S1**). Compared to the chronic stage, patients at 3 months follow-up (n=33 for both observations) recovered in the motor domain (left: p=0.046; right: p=0.002), in lateralized visual attention (p=0.030), while language-verbal memory and visuospatial memory-attention factors improved non significantly (**Supplementary Fig. S1**).

**Figure 3.**
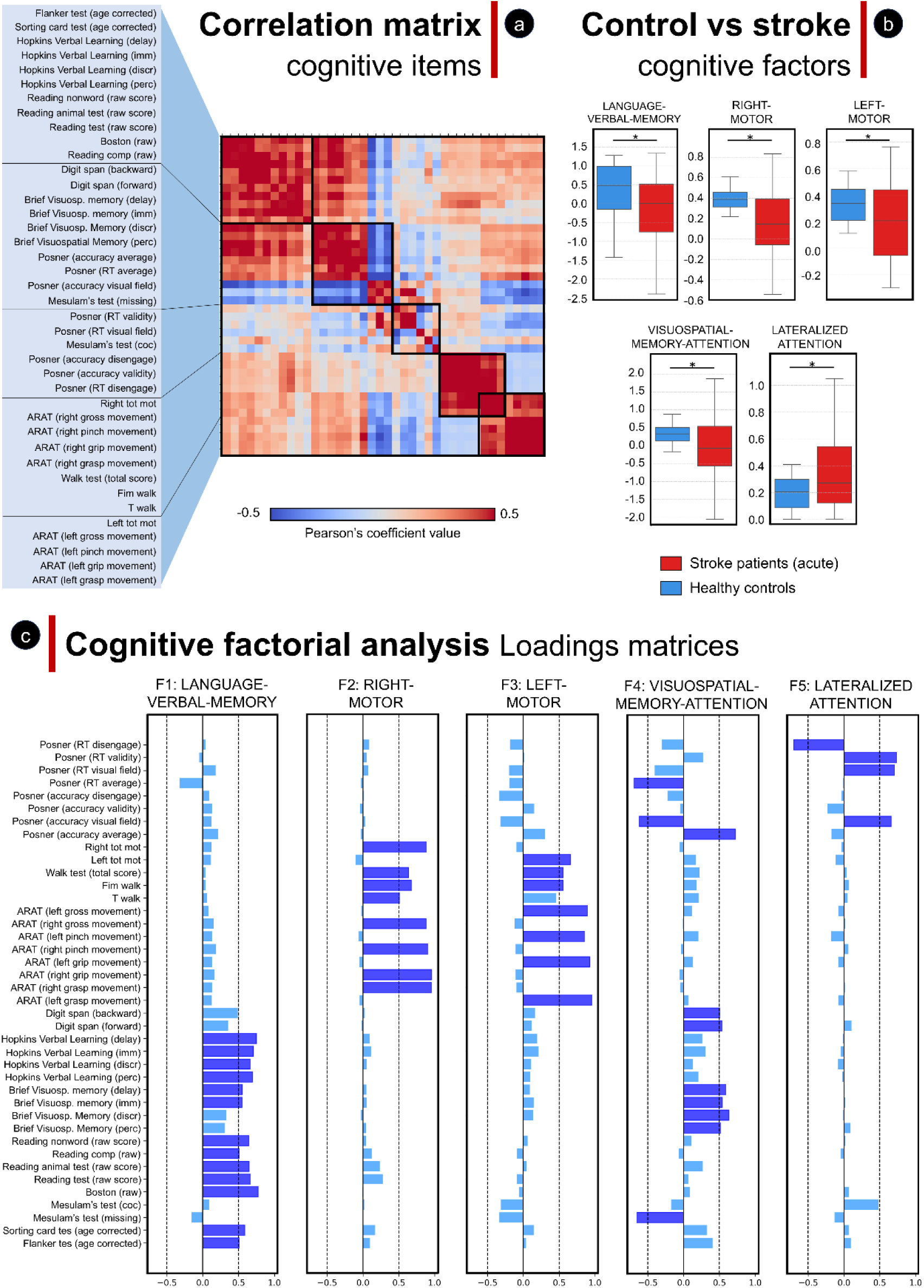
Latent factors of cognitive score. **Panel A**: Correlation matrix across all behavioral scores (a full description of the tests is reported in the Supplementary Materials). **Panel B**: differences between stroke at the acute stage (red) and controls (blue; visit 1) for the behavioral factorial scores. Significant differences are marked with *. **Panel C**: The five latent factors loadings for each behavioral test included in the analysis in panel B. In dark blue are highlighted the loadings with values higher than 0.5.

### Global disconnectivity patterns

The anatomical quantification of the SC gradients in HC was assessed both qualitatively and quantitively, showing that the identified structural latent patterns reflect the primary axes of WM fiber organization, capturing key variability along postero-anterior, ventral-dorsal, and left-right (callosal) directions. Specifically, in HC, for the intra-hemispheric connectivity the first gradient (_intra_G1) explained 22% of variance, the second gradient (_intra_G2) explained 17%, and the third gradient (_intra_G3) explained around 13% of variance. For the inter-hemispheric connectivity, _inter_G1, _inter_G2, and _inter_G3 explained 32%, 24%, and 15% of the variance, respectively. See **Fig. 2** for an overview of the gradients, **Supplementary materials S2.1** for an anatomical description, and **Supplementary Fig. S2** for the parcel-wise top 10% gradient values. The spatial patterns of these gradients were confirmed in controls at the first and second sessions (**Supplementary Fig. S2**). For the inter-hemispheric analysis n=4 patients were excluded due to at least a single parcel in the connectivity matrix with no values, not compatible with the gradient algorithm computation. Similar average-group patterns were found between stroke (sub-acute) and controls highlighting that gradients values captured similar connectivity axes in the two groups (**Supplementary Fig. S3**). The relationship between _intra_G1 and _intra_G2 highlighted a SC pattern moving from a posterior-anterior axis (highest _intra_G1 values), transitioning to a superior-inferior pattern (lowest _intra_G2 values), and returning to an anterior-posterior axis, following a ‘virtual’ U-shaped trajectory towards a positive relationship between _inter_G2 and _inter_G1 values (**Fig. 4-A**). The inter-connectivity relationship seems to capture corpus callosum anatomy moving from the anterior (highest _inter_G1 values) to the posterior (lowest _inter_G1 values) portion (**Fig. 4-B-C**).

**Figure 4.**
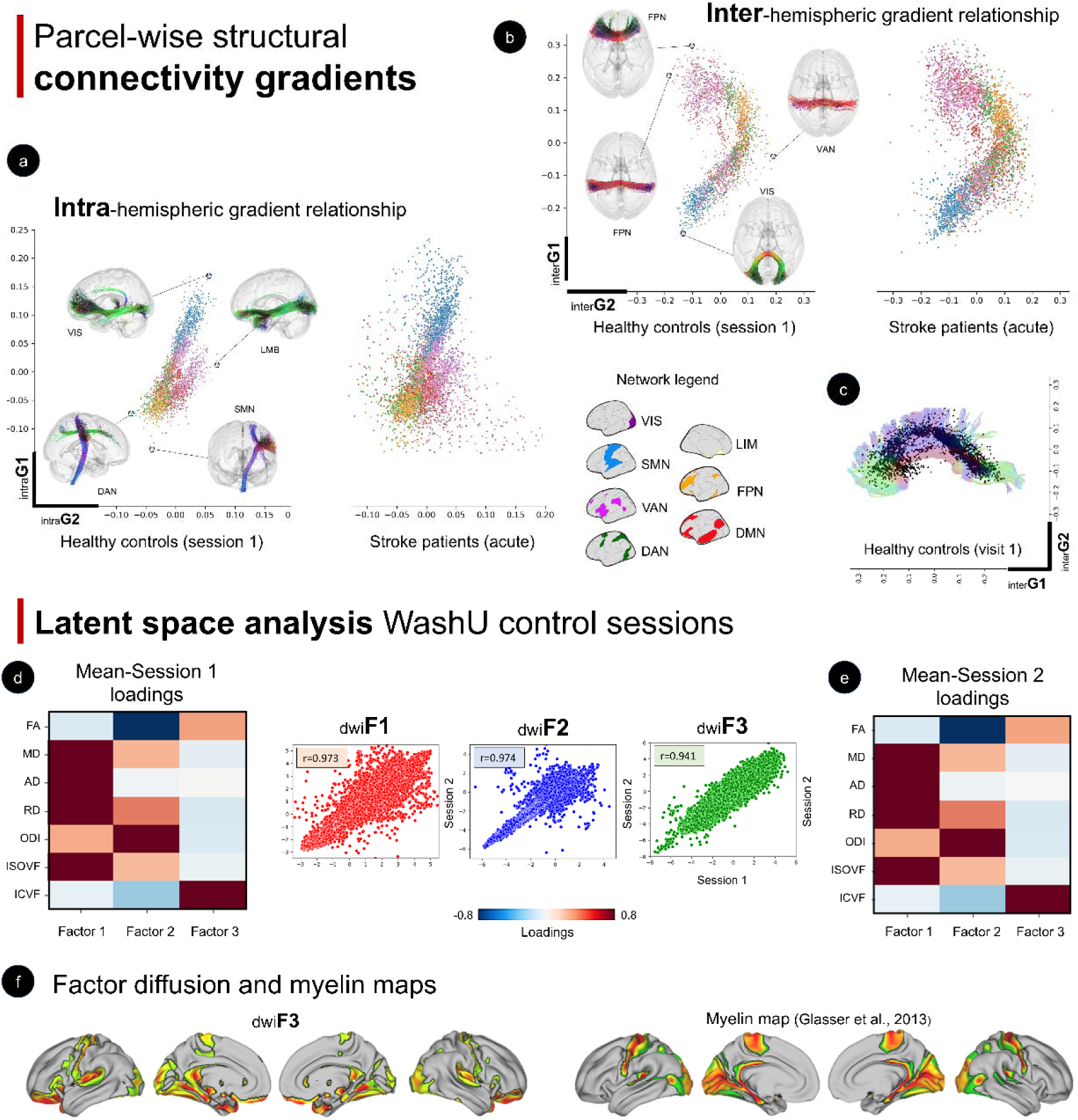
Structural connectivity gradients and microstructural factorial latent space. **Panel A**: Main gradient relationships in controls (left) and stroke patients (right) for intra-hemispheric connectivity; **Panel B**: Main gradient relationships in controls (left) and stroke patients (right) for intra-hemispheric connectivity. For each hemisphere, the interactions between G1 and G2 are presented. Exemplary tracts are reported for the extreme points in the cartesian axis (left, right, up, bottom). **Panel C**: Corpus callosum from the average template of the HCP-842 atlas (Yeh et al., 2018) was superimposed to the parcel-gradient values across the whole cohort of healthy controls from the WashU cohort. The relationship between interG1 and interG2 describes a spatial shape overlapping with the spatial topography of the corpus callosum (showed in partial transparency behind the black dots. The aim of this figure is to enhance the interpretability of the inter-hemispheric structural gradients. Abbreviations: DMN - default mode network; DAN - dorsal-attention network; SMN - sensorimotor network; LIM - limbic network; VIS - visual network; FPN - frontoparietal network; VAN - ventral-attention network. **Panel D**: microstructural factorial analysis results in the WashU controls cohort for the first MRI session; **Panel E**: microstructural factorial analysis results in the WashU controls cohort for the second MRI session. The scatterplots (center panels) highlight the spatial correlation between the weights projected at voxel level between the two sessions. **Panel F**: Vis-à-vis comparison between the third latent microstructural factor (_dwi_F3) from the HCP cohort and myelin maps from Glasser et al., (2013). Maps were thresholded to retain 94% of the highest values mapping to the cortex.

For the cross-sectional differences between controls and sub-acute stroke, first, we assessed whether the distribution of the relationship between the main gradients was similar between patients and controls. As shown in **Fig. 4**, the pattern, qualitatively, was similar between cohorts, although the former showed a more widespread distribution. SEM results showed significant differences for _intra_G1 and _intra_G2 mapping to SMN (Z=−2.13; p=0.033), DAN (Z=−2.68; p=0.007), and DMN (Z=3.07; p=0.002). The remaining networks were not significant (VAN: Z=−1.60; p=0.109; LIM: Z=−1.53; 0.125) or showed a trend towards significance (VIS: Z=1.92, p=0.055; FPN: Z=−1.94; p=0.052). For _inter_G1 and _inter_G2, sub-acute stroke patients and controls differed for the relationship within the VIS (Z=−8.76; p<0.001), SMN (Z=−3.27; p=0.001), DAN (Z=−4.78; p<0.001), VAN (Z=−4.99; p<0.001), and LIM (Z=−3.58; p<0.001). DMN and FPN showed no significant results (p>0.6). Second, the MANCOVA analysis on the UMAP space was conducted to assess these differences in the 2D-space (UMAP projection of parcel × gradient space). We found significant differences between sub-acute stroke and controls for all the four outcomes assessed for both intra-connectivity (ipsilesional hemisphere: F=6.70, p=0.0022; contralesional: F=3.43, p=0.0376) and inter-connectivity (ipsilesional: F=8.51, p=0.0005; contralesional: F=4.01, p=0.0225). Further, we assess the classification accuracy of this 2D space segregating controls and stroke by means of a SVM model. SVM, when applied to predict stroke and HC data points using UMAP space pooling contralesional and ipsilesional data computed from inter-connectivity gradients, reached a classification accuracy in the test set of 89% (**Fig 5-A**). When the SVM model was applied to intra-connectivity gradients we reached a classification accuracy of 96% in the test set (**Fig 5-A**). Details for the SVM hyperparameters validation are reported in the **Supplementary material S2.2**. Further, we performed a finer analysis at parcel-levels, assessing differences between stroke and controls by means of a one-sample t-test applied on GD values. Results confirmed the widespread differences at cortical levels, with the highest t-score widely distributed among the cortical parcels (**Fig 5-B**). These results were confirmed at the network-level analyzing directly gradient values (**Supplementary material S2.3** and **Supplementary Fig. S4**).

**Figure 5.**
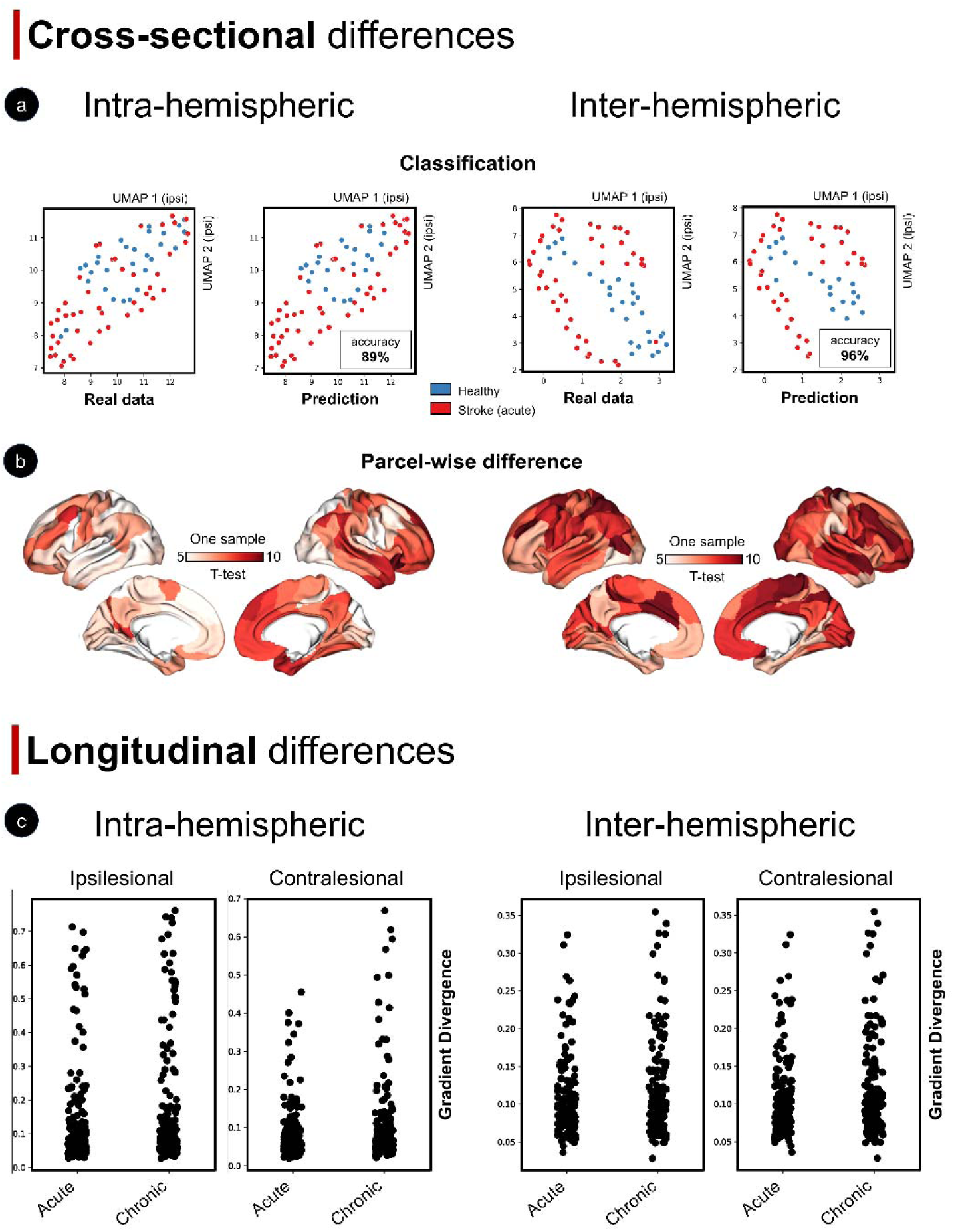
Cross-sectional and longitudinal structural connectivity gradient organization in stroke. **Panel A**: intra- and inter-hemispheric connectivity classifications (top panels) and parcel-level one-sample t-tests (bottom panels) are shown. The classification panels display the n^th^-dimensional gradient space projected into a 2D space, grouped by diagnostic labels (red: acute stroke; blue: controls – first session). The left plots show the grouping of the real data, while the right panels show the support vector machine classification accuracy for the same data distribution (accuracy is reported in the bottom box embedded in the plots). **Panel B:** parcel-wise one-sample t-test highlights parcels with the highest values (dark red), indicating greater deviation of stroke data from the control template in terms of gradient divergence. **Panel C**: Longitudinal changes in the PPA-L sample. Each dot represents the gradient divergence value at the network level for each individual included in the analysis. Networks are colored according to the scheme shown at the bottom, represented on the surface brain. Abbreviations: VIS: visual network; SMN: sensorimotor network; VAN: ventral attentional network; DAN: dorsal attentional network; LIM: limbic network; FPN: frontoparietal network; DMN: default mode network.

Subsequently, we quantified the longitudinal differences in stroke, comparing sub-acute (2 weeks) and chronic follow-up (3 months). This analysis was performed both in the whole sample (ITT-L) and reproduced in the subset of patients with complete longitudinal data (PPA-L). For the ITT-L sample, LMM analysis for the ipsilesional connectivity between sub-acute and chronic stage in stroke patients showed significant time effect for both _intra_GD (T=3.406; p=0.001) and _inter_GD (T=3.035; p=0.003). A significant effect was observed also when considering GD values for the contralesional hemisphere (_intra_GD, T=3.431, p=0.001; _inter_GD; T=2.667, p=0.008). Specifically, GD increased over time, suggesting an increase in the difference of the connectivity patterns with the controls’ template (**Fig 5-C**; **Supplementary Table S1-S2**).

### Local disconnectivity patterns

#### Microstructural factorial space description

DTI and NODDI maps from the two sessions of the HC from the WashU showed high reproducibility (**Supplementary Fig. S5-A**). From these maps, we reported 3 main latent diffusion factors with peculiar features in the cohort of HC from the WashU. The first factor (_dwi_F1) loaded on DTI diffusivity metrics (AD, RD, and MD) and ISOVF from NODDI. This factor explained around 52% of the variance. Anatomically, higher values were predominantly observed in the CSF, aligning with the loadings on diffusion metrics. The second factor (_dwi_F2) loaded on FA and ODI, with opposite directions, indicating a latent space capturing diffusivity orientation explained around 28% of the variance. Indeed, anatomically, divergent values were expressed in the GM and WM tissues, the latter showing lower values (thus higher anisotropy in the WM). The third factor (_dwi_F3) loaded on both FA and ICVF, along the same direction, explaining 15% of the variance of the data (**Fig. 4-D-E**). This last factor showed relative spatial similarity with the myelin map distribution from Glasser et al. (2016), suggesting a microstructural pattern following the main (and myelinated) tracts (**Fig. 4-F**). The latent microstructural space reported in the second session of the cohort of HC from WashU was used to project microstructural data of stroke patients. After this transformation, four individuals were excluded from the subsequent analysis due to differences with the control cohort exceeding an arbitrary threshold of similarity with the template (spatial correlation r<0.2) for the _dwi_F1. Although visual quality control showed similar diffusion factor patterns, these patients were removed to avoid potential biases in the microstructural analysis. An additional analysis computing the latent space independently on the stroke cohort at the acute stage and 3 months, echoed the findings in the HC (**Supplementary Fig. S5-B**). These results were in line with the findings on the HCP dataset (**Supplementary Material S2.4**) and both at group level and single subjects’ space (**Supplementary Fig. S6**). Moreover, factors were highly reproducible across the two independent datasets (WashU and HCP), with an average correlation above 0.9 for loadings patterns (F1: sess1-sess2 r=0.973, WashU-HCP r=0.967; F2: sess1-sess2 r=0.974, WashU-HCP r=0.994; F3: sess1-sess2 r=0.941, WashU-HCP r=0.939) (**Supplementary Fig. S6**).

Two main structural disconnection patterns were observed running a descriptive PCA on the SDC maps, representing more than 66% of the variance. These components mapped within the left and the right hemisphere, respectively (**Fig. 2**). In line with the lesion frequency map (**Fig. 2**) the right disconnection pattern captured 42% of the variance, compared to the left disconnection pattern (23%). SDC maps from individual lesions were used as masks for the latent microstructural parameters analysis. SDC maps showed significant differences comparing stroke acutely with controls for the second factor (_dwi_F2) when considering a probability disconnection of 60% (U=1277; p=0.033) and 80% (U=1135; p=0.024), while the 40% threshold showed a trend towards significance (p=0.06). The remaining factors were not significantly different from stroke and controls (p>0.6)(**Fig. 6-A**). A significant involvement of the second factor was confirmed at the voxel-wise level. Higher _dwi_F2 values were observed in stroke patients acutely compared to controls (p<0.05 FWE-corrected), while no significant voxels were reported for _dwi_F1 and _dwi_F3 (**Supplementary Material S2.5** and **Supplementary Fig. S7**). Higher _dwi_F2 values in stroke are indicative of lower anisotropy in the WM, consistent with reduced WM integrity and the main involvement of the subcortical WM in stroke and related disconnection ^26,51,53^. Next, we were interested in measuring the effect of disconnection on the contralesional GM. Accordingly, we ran a linear regression analysis including microstructural GM contralesional values from _dwi_F1, _dwi_F2, and _dwi_F3 as independent variables and structural GM contralesional probability disconnection as the dependent variable. We found significant results at the sub-acute stage (_adj_r2=0.625; p<0.0001) for both _dwi_F2 (coef:-0.244; p<0.001) and _dwi_F3 (coef:0.132; p=0.012). Higher disconnectivity probability was accompanied by lower _dwi_F2 and higher _dwi_F3, suggestive of pathological mechanisms in the connected GM tissue (e.g., neuroinflammation and decreased dendrite density/arborizations)^37^. Results were echoed by the bootstrapping procedure that showed a significant effect for the structural model (F=21.65, p<0.0001)(**Fig. 6-B**). This result indicates a significant linear relationship between the degree of WM disconnection and the alteration of the diffusion properties of the contralesional GM hemisphere. To confirm these findings, we ran a separate analysis in which we compared parcels with the highest and the lowest probability of disconnections. A significant effect was found for _dwi_F2 (**Fig. 6-C**). The result was highly stable across the random selection of parcels with the lowest probability (n=500). Moreover, _dwi_F2 showed the highest differences between parcels compared to _dwi_F1 and _dwi_F3 assessed with a one-way ANOVA (F=347, p<0.00001; np^2^=0.32). The results were replicated when excluding the subcortical parcels from the analysis (F=26.3, p<0.00001; np^2^=0.15), controlling for lesion location (**Supplementary Fig. S8**).

**Figure 6.**
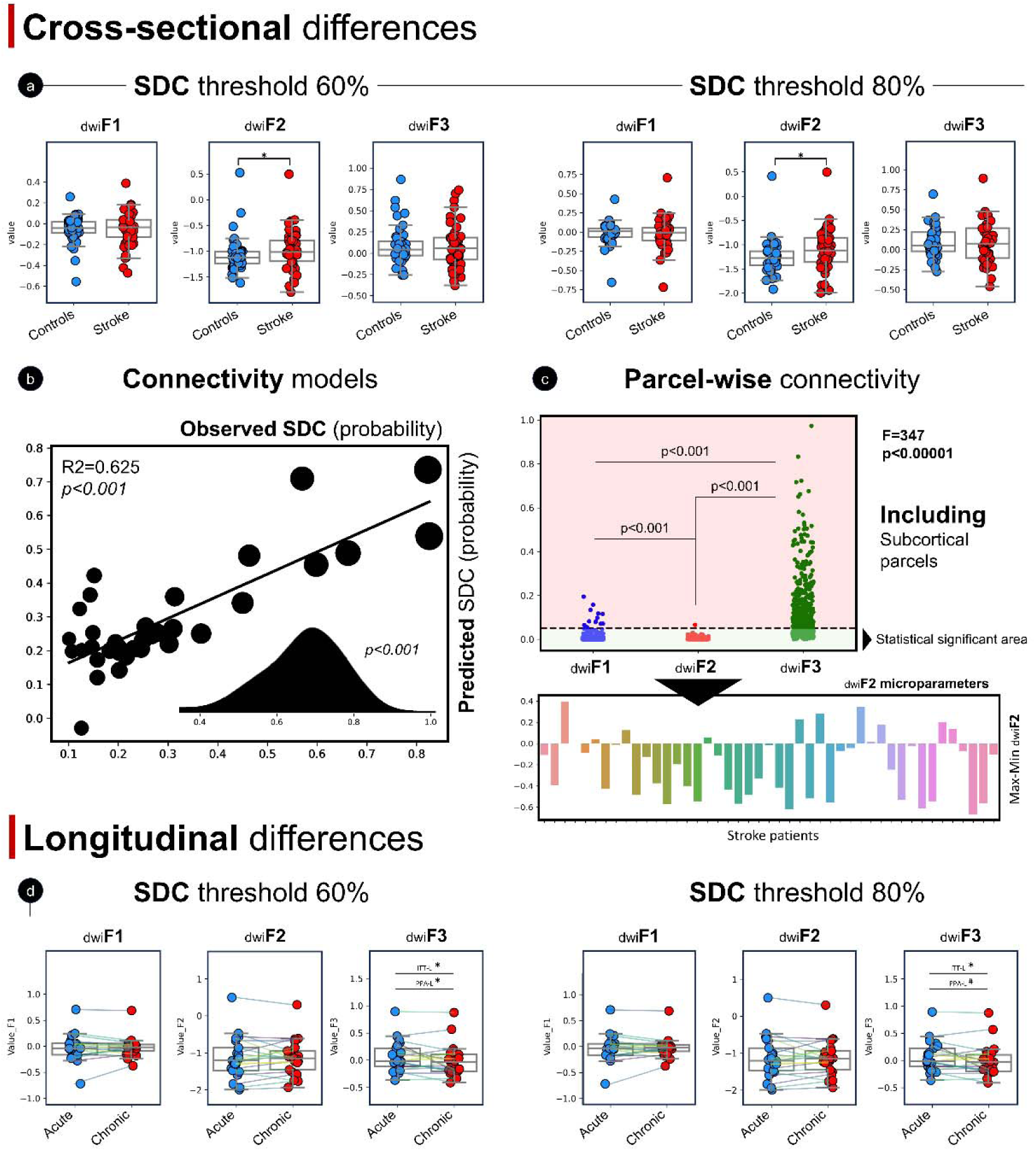
Cross-sectional and longitudinal microstructural latent factors in stroke. **Panel a**: cross-sectional differences in microstructural latent factors between stroke (acute) and controls. Each plot represents lesion values for patients (red dots) and the values associated with the lesion location in the controls (blue dots). Results are presented for all microstructural factors and two different SDC thresholds. Asterisks (*) indicate significant p-values. linear model illustrating the relationship between microstructural properties and the dysconnectivity profile of contralesional regions during the acute stage. The scatterplot displays observed versus predicted values from the linear model (values are coded according. The distribution of R² values from the bootstrap procedure is shown in the subpanel in the top-left corner. **Panel c**: microstructural latent properties within parcels classified as disconnected *versus* not disconnected from the lesion. The procedure was iteratively repeated by randomly selecting the least disconnected parcel. Each plot in the panel shows the p-value associated with each iteration. Significant p-values are in the bottom region (green), while non-significant p-values are in the top (red) zone. The bar plot illustrates the difference in _dwi_F2 values (y-axis) between the maximally disconnected parcel and the iteration-average minimally disconnected parcel for each patient included in the analysis (x-axis). **Panel D**: Longitudinal differences in microstructural latent factors between the acute and chronic stages. Significance is reported for both the ITT-L and PPA-L samples. Asterisks (*) indicate significant p-values, while hash marks (#) indicate trends toward significance.

Longitudinal changes in latent microstructural parameters within SDC masks using LMM were significant for _dwi_F3 at different disconnectivity thresholds (SDC-40%: T=2.124, CI[2.5-97.5]=0.113-0.752, p=0.043; SDC-60%: T=2.747, CI[2.5-97.5]=0.022-0.133, p=0.011; SDC-80%: T=2.128, CI[2.5-97.5]=0.006-0.150, p=0.043). Specifically, we found that SDC regions (independently by the threshold applied) showed on average lower values at the chronic stage compared to the sub-acute stage, suggestive of longitudinal degeneration (**Fig.6-D**). Results were confirmed when including the PPA-L cohort considering both the gradients (n=26 stroke patients for intra-connectivity and n=22 for inter-connectivity analysis) and the latent microstructural space (**Supplementary Table S1-S2**). Additionally, a post-hoc sensitivity analysis was conducted, focusing exclusively on the SDC maps, to evaluate whether the latent factor computed at the individual level (without projection into the HC-session space) would yield similar results. The analysis on the ITT-L sample demonstrated stable and consistent findings, with significant changes in the same direction appearing only in _dwi_F3 across all probability thresholds (SDC-40%: T=3.686, CI[2.5-97.5]=0.027-0.152, p=0.001; SDC-60%: T=4.062, CI[2.5-97.5]=0.029-0.176, p<0.001; SDC-80%: T=3.131, CI[2.5-97.5]=0.039-0.199, p=0.005). No significant differences emerged considering _dwi_F1 and _dwi_F2.

### Correlation with behavior

We assessed the linear relationship between SC gradients and behavior. At the sub-acute stage, stroke-related changes in intra-hemispheric structural gradients were significantly associated with deficits in language-verbal memory, visuospatial and spatial memory, and attention-neglect (**Fig 7-A**). Language-verbal-memory deficits mapped to perisylvian, prefrontal, and parietal cortices of the left hemisphere, while both visuospatial memory and attention/neglect functions mapped widely to prefrontal, parietal, and temporal regions of both hemispheres, more strongly on the right hemisphere. Visuospatial memory and attention/neglect deficits mapped significantly to inter-hemispheric structural gradients, consistent with a view of inter-hemispheric competition for spatial attention.^72^ In contrast, motor deficits were not significantly associated with changes in structural gradients (**Fig 7-A**). All the results were FDR-corrected (p<0.05). At the chronic stage, we found no significant behavioral association for the intra-hemispheric GD. A weak association for the visuospatial-memory-attention and the lateralized-neglect components was reported, mapping mainly in the parietal and occipital regions of the left hemisphere. Data reporting significant correlations for both sub-acute and chronic follow-up are shown in the **Supplementary Fig. S9**.

**Figure 7.**
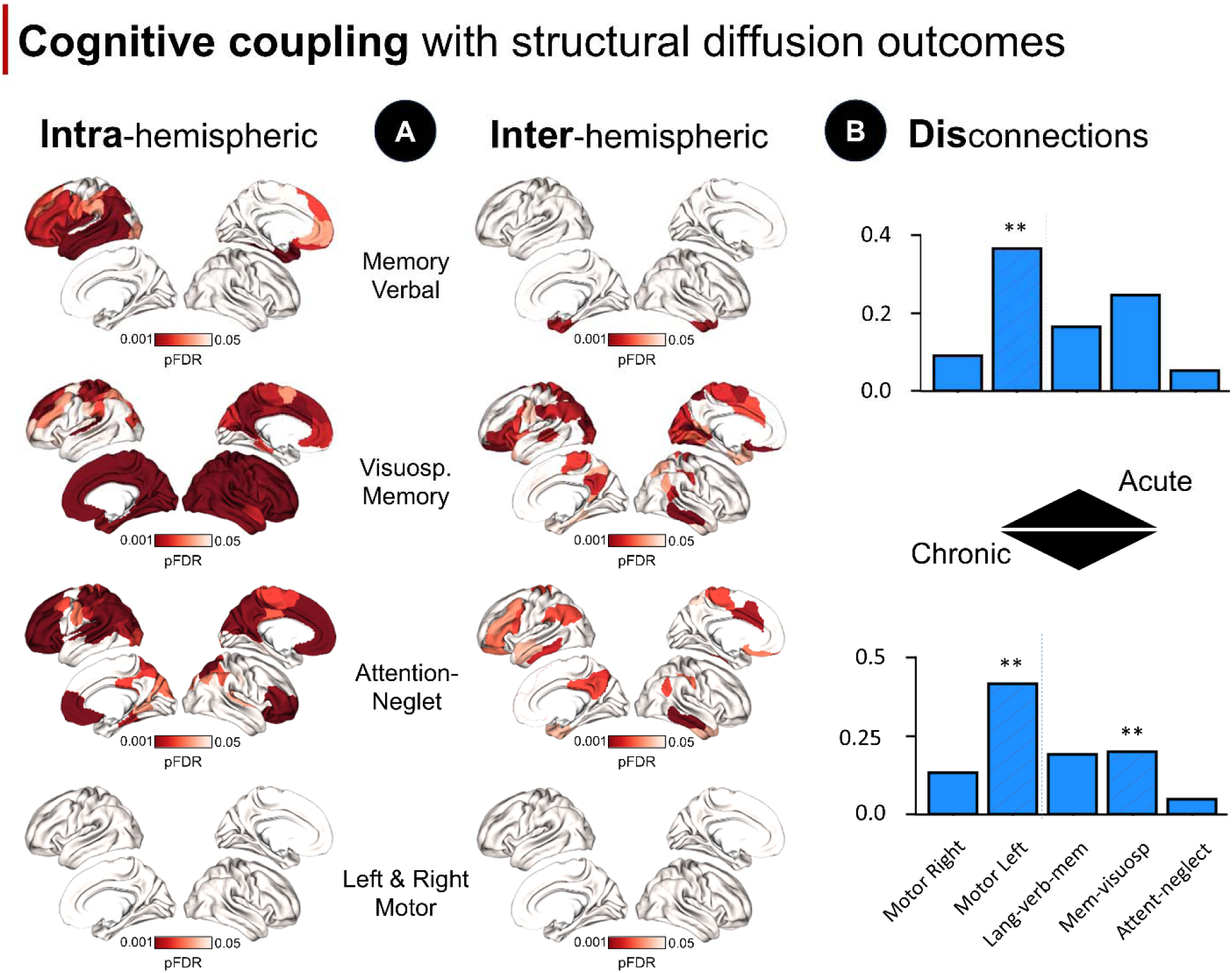
Cognitive relationship with structural connectivity patterns and microstructural factors. **Panel A**: Parcel-wise cognitive-gradient correlation at the sub-acute stroke stage. The linear correlation between behavioral scores and GD values at the parcel level was computed for each behavioral factor. Only parcels surviving p<0.05 FDR correction are shown. Results are reported for both intra-(left panels) and inter-(right panels) connectivity gradients. The chronic stage showed marginal significance, and results are reported in **Supplementary Figures S9**. **Panel B**: Microstructural latent space and behavioral deficits at acute and chronic stages. Top panels: linear relationship between behavioral factors and the microstructural factors within the region-of-interest considered by the study (disconnectivity maps) at the acute stage. Each boxplot represents the r^2^ of the linear model predicting behavioral scores by means of the latent microstructural values. Bottom panel: the same analysis was repeated at the chronic stage. ** marks p-values < 0.01 surviving multiple comparison significance.

In contrast to the structural gradients, a significant relationship occurred between motor left deficits and SDC surviving multiple comparison correction (r^2^=0.37; *p=0.003*). The remaining behavioral factors did not show significant associations (**Fig. 7-B**). The relationship between SDC microparameters and left motor deficits remained significant at the chronic stage (left: r^2^=0.415; *p<0.001*), with right motor scores showing a trend towards significance (r^2^=0.133; p=0.058)(**Fig. 7-B**). Further, a significant association emerged within the SDC maps the visuospatial-memory-attention components not surviving the significant threshold (r^2^=0.25; p=0.046), while the association with the language-verbal-memory component survived the statistical threshold (r^2^=0.190; *p=0.008*).

### Structural gradient and local disconnectivity association

For this analysis, only _dwi_F2 and _dwi_F3 were considered, given the marginal contribution of _dwi_F1 to the results. Microstructural outcomes within the SDC mask (40% cut-off) were significantly predictive of the global GD value across all comparisons considered: intra-hemispheric contralesional (R² = 0.186, p = 0.020), intra-hemispheric ipsilesional (R² = 0.224, p = 0.008), inter-hemispheric contralesional (R² = 0.228, p = 0.011), and inter-hemispheric ipsilesional (R² = 0.167, p = 0.041). Specifically, as GD increased, in the disconnected WM tracts _dwi_F2 increased, while _dwi_F3 decreased, indicating axonal disorganization/degeneration due to increased extra-axonal space.^37^ Detailed results are provided in **Supplementary Fig. S10** and **Supplementary Table S3.** At the network level, these results were confirmed (except for inter-hemispheric ipsilesional connectivity), showing a trend involving most of the networks considered. This is consistent with the GD results, which indicate a global network pattern (**Supplementary Fig. S10**).

### Sensitivity analysis comparing results from cortical and subcortical lesions

Sensitivity analysis largely confirmed the above results when comparing cortical and subcortical lesions, suggesting robust gradient alterations irrespective of lesion location (**Supplementary Material S2.6 and Fig. S11**).

## 4. DISCUSSION

This is the first study that combines large-scale gradient mapping and microstructural analysis of disconnection in stroke. We implemented novel methods to compute gradients of SC across the brain, which can capture the similarity of tractography-derived connectivity patterns between brain regions. We replicated our results in a robust structural gradient organization in a large dataset of controls (HCP) and demonstrated how focal stroke lesions widely affect this organization and worsen longitudinally. For local microstructural analysis, we applied novel embedding methods to extract latent variables that capture variability across both DTI and NODDI measures, describing microstructural properties voxel-wise. This approach was validated in two independent cohorts and demonstrated longitudinal changes in stroke patients, coupled with structural gradient changes. Finally, we demonstrated how divergence in SC gradients relates to underlying microstructural changes, providing a mechanistic link between global network reorganization and local tissue properties.

We applied the framework of gradient mapping,^73^ to high quality dMRI data. The three gradient maps summarize the main axes of structural organization, in line with previous studies using the same methods,^61,74,75^ while only partially overlap with functional gradient maps derived from fMRI.^61,73,74^ In stroke, the gradient analysis showed widespread differences at the sub-acute stage, as compared to controls, both in intra-hemispheric connectivity, ipsilesional and contralesional to the lesion, and inter-hemispheric connectivity. The abnormalities were widespread across all networks, in line with previous studies that found global alterations of SC after focal stroke lesions,^33,39,40^ and intra-hemispheric and inter-hemispheric changes in functional networks post-stroke.^15,23,76,77^ The structural changes were large enough that a stroke patient could be distinguished from HC at the single subject level with an accuracy of around 90%. When the structural gradients were re-measured at 3 months post-stroke we showed a growing deviation from the HC’s template. This is interesting as local synaptic sprouting and remodeling of the WM have been proposed as a mechanism of recovery in animal models,^57,78^ and most recovery has already occurred at three months.^79^ However, our results are in line with other human studies showing progressive structural degeneration following stroke.^30,41,43^ The structural degeneration of WM connections is in stark contrast with functional data showing a normalization of FC correlating with recovery of function.^24,48^ Since at the sub-acute stage, changes in FC are closely aligned with alterations of SC,^26^ it derives in the course of recovery structural and FC trajectories diverge, with structure degenerating and function normalizing likely based on synaptic reweighting of surviving connections. This adaptive capacity for FC might drive behavioral recovery especially in more redundant cognitive networks where the link between structure and function is weaker^74^ (see below behavioral correlation).

One of the most influential results in the last decade has been realizing the importance of disconnection on brain networks caused by focal lesions.^23,76^ Recent methods have been developed to infer (*indirectly*) patterns of functional and structural disconnection based on normative atlases.^50,80^ Here we combine for the first-time *direct* measures of diffusivity microstructure to the analysis of disconnected brain regions after focal stroke lesions and relate these changes to global changes of connectivity measured with the structural gradients.

First, we described a latent microstructural space showing high reliability in condensing complex diffusion metrics into a few stable and interpretable components. Three latent diffusion factors were identified robustly in both HCP and WashU cohorts, with high replicability across sessions and participants. Next, we mapped these latent diffusion microparameters to gray and WM regions identified as disconnected.^50^ We found at the sub-acute stage disorganization in disconnected WM and disconnected cortical parcels (mapping on _dwi_F2 values). This pattern is consistent with decreased FA in WM, hence fiber disorganization, and decreased ODI in the GM, hence loss of neurite dispersion suggestive of cortical deafferentation. Hence disconnected regions show sub-acutely a loss of long-range WM pathways and an impact on the corresponding homotopic contralesional cortical regions. Interestingly, these disconnected regions further degenerate from 2 weeks to 3 months as indicated by a decrease of _dwi_F3, a measure indicative of myelination.^81^ Therefore in line with a previous study,^30^ WM fibers and gray matter disconnected from the lesion showed degeneration over time post-stroke.

Interestingly, the changes of microparameters in disconnected tissue partially but significantly explained the SC distance between stroke patients and HC. Higher _dwi_F2 and lower _dwi_F3 predicted GD values in stroke, suggesting a complex relationship between structural gradient divergence and microstructural alterations. While this result is trivial for large lesions that disconnect large parts of the brain, it is less clear for small lesions that produce smaller maps of disconnection, yet apparently still produce large alterations of large-scale structural organization. In previous work we have estimated that nearly 20% of all region pairs were estimated to be either directly or indirectly disconnected by stroke lesions in our sample, with extensive disconnections associated primarily with damage to deep WM locations.^26,54^

We measured behavior using a large battery, finding five factors that explained about 50% of the behavioral variance across groups and time points. This low dimensionality is consistent with the factorial-structure found in stroke,^82,58^ and in other conditions such as brain tumors^83^ and neurodegeneration.^59^ While in stroke they likely reflect both the correlated topography of vascular lesions^84^ and the low dimensionality of FC abnormalities,^15,20^ more generally they are thought to reflect loss of neural entropy^85^ caused by any neurological disorder.

Interestingly, different behavioral deficits showed a dissociated pattern of behavioral correlation with SC. Cognitive functions robustly correlated at the sub-acute stage with structural gradients. The topography was highly specific with left hemisphere alterations related to verbal-language-memory, bilateral alterations for visuospatial-memory-attention and lateralized attention. Interestingly, motor deficits did not correlate with changes in structural organization. This result is in line with previous studies in which distributed intra- and inter-hemispheric FC patterns correlated with sub-acute deficits in language, attention, and memory, but not visual or motor deficits^20^ (as noted, the current work was based on an independent sample than our previously published stroke papers. Moreover, imaging data was collected using a different scanner and for behavioral data was used a modified battery that eliminated some subtests and added others). Structural gradient changes at 3 months were weakly related with cognitive tasks, which indicates a decoupling between SC and cognitive performance. Given normalization of FC correlates with the behavioral recovery of cognitive functions,^48^ our results show that behavioral recovery depends on synaptic reweighting of residual structural connections in the context of longitudinal structural degeneration. This view is also consistent with recent work in non-human primates highlighting the role of transmodal association cortex in post-stroke recovery.^24^

In contrast to the cognitive domains, motor deficits (sub-acute and chronic stages) correlated with microstructural parameters of disconnected regions. Since the lesion distribution was mainly located in the central WM underlying brain motor regions, and the main patterns of disconnection were correspondingly centered around the central sulcus including the corticospinal tracts, the association between motor deficits and the microstructure of disconnected regions indicates the importance of local damage/disconnection in accounting for motor and sensory deficits in line with our previous work.^20,48,51,54,58^

This study is not without limitations. Gradient mapping is sensitive to methodological choices, which can affect the reproducibility and comparability of results. Further, the interpretation of gradients is more intricate compared to more established methods, including the lack of consensus for interpreting gradient results. Despite these limitations, several strengths should be highlighted. The use of multi-shell diffusion data has significantly improved the tractography, contributing to the reliability of our findings. The longitudinal aspect of our data collection has provided valuable insights into the temporal evolution of stroke. Moreover, the high-level analysis that integrates microstructural, structural organization and behavioral data has unveiled latent patterns and relationships that may not be apparent through traditional methods.

## Data Availability

All data produced in the present study are available upon reasonable request to the authors.

## Acknowledgements

We wish to thank the participants who participated in the study for their time.

## Author contribution

LP: methodology, design, conceptualization, formal analysis, statistics, interpretation, data visualization and writing of the draft; MHA: methodology, design, conceptualization, formal analysis, interpretation, and writing of the draft; AS: revising the draft; AMC: Washington University stroke database; NVM: Washington University stroke database; JCG: Washington University stroke database; ARC: Washington University stroke database; GLS: Washington University stroke database; MC: data interpretation, design, conceptualization, supervision, and revising the draft.

