## Supplementary materials for "Longitudinal degeneration of microstructural and structural connectivity patterns following stroke"

Number of words supplementary text: 3294

Number of figures: 10

Number of tables: 2

***Corresponding author:**

Prof. Maurizio Corbetta, Clinica Neurologica,

Department of Neuroscience, University of Padova, via Giustiniani 2, Padova 35131, Italy.

University of Padova

**S1. SUPPLEMENTARY MATERIALS**

**S1.1. Inclusion and exclusion criteria**

Inclusion criteria were: 1) first-time symptomatic ischemic or hemorrhagic stroke; 2) enrollment within 1-3 weeks of stroke onset; 3) age of 18 or higher; 4) no more than two lacunes, with less than 15 mm size on CT scan and no symptoms; 5) clinical signs of visual, motor, language, attention, or memory impairment on examination; 6) alertness, wakefulness, and ability to participate in the study. We excluded patients i) with prior stroke or multifocal lesions according to clinical imaging, ii) inability to remain awake during testing, iii) patients with other relevant neurological, psychiatric, or medical conditions that could interfere with the participation to the study, iv) patients with less than one-year life expectancy (e.g., cancer or congestive heart failure class IV).

**S1.2. Behavioral tests**

*S1.2.1. Motor Battery*

1) Functional performance with the Action Research Arm Test (ARAT), evaluating grasp, grip, pinch, and gross motor movements (Van Der Lee et al., 2001). 2) Combined Walking Index, timing patients' 10-meter walk or using the Functional Independence Measure's Walking item for those unable to walk independently. The following scores were computed: i) timed walk for 10 meters (*T Walk* in Figure 3 and Figure S1); ii) functional Independence measure, a measure assessing assistance levels for walking (*Fim* in Figure 3 and Figure S1), from 1 (total assistance) to 7 (independent walking); iii) a combined score (timed walk / functional independence measure, *Walk test total score* in Figure 3 and Figure S1) (Keith, RA et al., 1987; Kempen et al., 2011; Perry et al., 1995). 3) Total Motricity Index (MI), adding muscle testing scores for hip flexion, knee extension, and ankle dorsiflexion on each side. For both left and right side of the body a total lower extremity MI was computed according to the following formula: ankle MI + knee MI + hip MI + 1 (*Left tot mot* – for left – *Right tot mot* – for right – in Figure 3 and Figure S1)

*S1.2.2. Memory*

1) Visual memory was assessed with the Brief Visuospatial Memory Test-Revised (BVMT-R) (Benedict, 1997). Patients viewed abstract figures and reproduced them from memory across three immediate recall trials and one delayed recall trial, followed by a recognition test. Scores included: i) immediate total recall T-score (age-normed corrected using normative data from the test manual; *Imm* in Figure 3 and Figure S1), ii) delayed recall T-score (age-normed; *Delay* in Figure 3 and Figure S1), iii) delayed recall percent retained (computed from the percent items retained from last immediate recall trial to delayed recall; *Perc* in Figure 3 and Figure S1), and iv) delayed recognition discrimination index (proportion of correct recognitions, correct rejections, misses, and false alarms, using the table provided in the test manual; *discr* in Figure 3 and Figure S1). 2) Verbal memory was evaluated with the Hopkins Verbal Learning Test-Revised (HVLT-R) (Benedict et al., 1998), where patients heard a word list and recalled it over three immediate trials and one delayed trial, followed by a recognition test. Scores included: i) immediate total recall T-score (age-normed corrected using normative data from the test manual; *Imm* in Figure 3 and Figure S1), ii) delayed recall T-score (age-normed; *Delay* in Figure 3 and Figure S1), iii) delayed recall percent retained (percent items retained from last immediate recall trial to delayed recall; *Perc* in Figure 3 and Figure S1), and iv) delayed recognition discrimination index (proportion of correct recognitions, correct rejections, misses, and false alarms, age-normed using the table provided in the test manual; *Discr* in Figure 3 and Figure S1). 3) Spatial working memory was tested with the Wechsler Memory Scale subtest (Delis et al., 2012). Patients copied sequences of taps on a block board and later reproduced them in reverse. Scores included: i) spatial span *Forward* and *Backwards*.

*S1.2.3. Language*

Battery Subtests of the Boston Diagnostic Aphasia Examination (BDAE-III) were performed according to the standard protocol (Roth, 2011): The core assessment comprised four language processing tasks: 1). Comprehension of Oral Reading of Sentences: Patients answered multiple-choice comprehension questions about the sentences they just read (*Reading comp* in Figure 3 and Figure S1); 2). Boston Naming Test short form: patients named the item pictured (*Boston* in Figure 3 and Figure S1); 3) Non-word Reading: Four-letter nonwords (e.g. NORD) were presented (*Reading nonword* in Figure 3 and Figure S1); 4). Oral Reading of Sentences: Patients read sentences aloud (*Reading test* in Figure 3 and Figure S1).

*S1.2.4. Executive functions*

A subtest of the Delis-Kaplan Executive Function System was performed according to the standard protocol: Animal Naming (*Reading animal test* in Figure 3 and Figure S1): patients named as many animals as possible in 1 minute (Tombaugh, 1999).

Two additional tests were included from the NIH Toolbox (Weintraub et al., 2013): 1) The Flanker score (age corrected; *Flanker test* in Figure 3 and Figure S1) which  tests the ability to inhibit visual attention to irrelevant task dimensions; 2) The Card sort test (age corrected; *Sorting card test* in Figure 3 and Figure S1) that assesses the set-shifting component of executive functions.

*S1.2.5. Attention*

Different visuospatial attention processes were measured with the Posner orienting task (Posner et al., 1984). The experimental setup consisted of an Apple Power Macintosh computer with a 17-inch Apple Monitor and a Carnegie Mellon button box for response collection. Eye movements were visually monitored, with fixation breaks promptly addressed. The display configuration featured a central fixation cross and two peripheral square frames (1 degree per side, centered 3.3 degrees from fixation) along the horizontal meridian. For patients with quadrantanopsia, stimuli were presented in visible field regions at symmetrically opposite positions across the vertical meridian. Each trial proceeded as follows: A red-to-green color change of the fixation cross initiated the trial, followed 800ms later by a directional arrow cue (left/right) at fixation, displayed for 2360ms. After a variable delay (1000-2000ms), a target asterisk appeared for 300ms within either frame. The target appeared at the cued location in 75% of trials (valid condition) and at the opposite location in 25% (invalid condition). Patients responded via key-press using their ipsilesional hand, with a 2360ms intertrial interval. Two 40-trial blocks (30 valid, 10 invalid) were administered, requiring 15 minutes including practice.

The assessment measured: 1). The reaction time (RT) validity effect: difference in reaction times between validly cued trials (where the cue correctly predicts the target's location) and invalidly cued trials (where the cue misdirects attention)(*Posner (RT validity)* in Figure 3 and Figure S1); 2) The RT disengagement subtest: RT when attention must be shifted away from an invalid cue to the actual target location (*Posner (RT disengage)* in Figure 3 and Figure S1); 3) The RT Visual Field effect: assesses differences in RT between targets presented in the left versus right visual field, indicating potential lateralized attentional biases (*Posner (RT visual field)* in Figure 3 and Figure S1). 4) The Accuracy disengagement: evaluates the reduction in accuracy when attention must be redirected from an invalid cue to the actual target location (*Posner (accuracy disengagement)* in Figure 3 and Figure S1); 5) The accuracy subtest: assesses difference in accuracy between validly cued trials (where the cue correctly predicts the target's location) and invalidly cued trials (where the cue misdirects attention).

Additionally, visuomotor spatial function was evaluated using a standardized instrument: the Mesulam Unstructured Symbol Cancellation Test. The Mesulam Center-of-cancellation score was calculated using Rorden and Karnath (2010) software to quantify lateralized miss patterns.

*S1.2.6. Factorial analysis*

A hierarchical factorial analysis was performed to reduce the dimensionality of behavioral data. Kaiser-Meyer-Olkin (KMO) was assessed for consistency of data with dimensionality reduction approach. Before running the analysis, the cognitive dataset was processed and cleaned according to following steps: patients with more than 15% of missing tests/subtests were excluded from the analysis. The remaining missing data were substituted with the group mean value, a standard procedure for dealing with missing values. Data were finally z-scored according to the mean and standard deviation of the whole cohort, including control subjects and stroke patients at both acute and chronic (3 months) stage. This procedure was preferred rather than computing latent factors in controls and projected to stroke patients due to: i) the relatively small sample size of controls; ii) ceiling effect in several tests.

All possible numbers of factors from 1 to the maximum number were examined according to three main criteria: (a) model fit, (b) interpretability, and (c) robustness. Model fit was measured as the percentage of total variance explained and should be as high as possible while still maintaining the other two criteria. For interpretability, we adopted the guidelines from Velicer and Fava (1998), suggesting that each factor should have at least three variables with loadings > 0.4. For enhancing robustness this threshold was heightened to 0.5. To ensure the robustness of this approach we computed the cognitive latent factors also in the cohort of stroke patients at the acute stage. Further, latent factors with eigenvalues higher than 1 and explaining more than 5% of variance were retained, in line with our previous studies (Corbetta et al., 2015; Pini et al., 2022).

**S1.3. Diffusion imaging preprocessing**

T1w weighted imaging data were normalized through non-linear approaches (FSL-FNIRT) to the MNI template. Lesions were manually segmented on the normalized structural MRI images using the Analyze biomedical imaging software system (Robb and Hanson, 1991). For DWI preprocessing, the PreQual pipeline (Cai et al., 2021) was applied to create a synthetic susceptibility-corrected b0 volume using SYNB0-DISCO to correct susceptibility-induced artifacts a deep learning framework by Schilling et al. (2019). This approach allows to correct for distortion correction without acquiring DWI images in the opposite phase-encoding direction but estimating the distortion from structural images. The synthetic b0 image was used within FSL's topup algorithm to correct susceptibility-induced artifacts in DWI data. Then, DWI raw data were denoised using the MP-Principal Component Analysis (PCA) function included with MRTrix3 (Veraart et al., 2016). The local subvoxel-shifts method (Kellner et al., 2016) was employed to reduce Gibbs ringing artifacts. Rician correction was performed with the moments method (Koay and Basser, 2006). Volumes with a corresponding b value under 50 were considered b0 volumes for the pipeline's remaining steps. FSL's eddy algorithm was used to correct for motion artifacts, eddy currents, and remove outlier slices (Andersson and Sotiropoulos, 2016). N4 bias field correction was applied to correct for bias inhomogeneity (Tustison et al., 2010). Lastly, the preprocessed data were fitted with a tensor model using MRTrix3's dwi2tensor function and an iterative reweighted least-squares estimator (Veraart et al., 2013). An extensive DWI preprocessing quality check was employed.

**S1.4. Structural connectivity computation: tractography and gradients**

This process involved skull stripping, segmentation, and tissue classification of T1w images, followed by non-linear registration and five-tissue-type segmentation for tractography. Cortical surface segmentations were done using FreeSurfer (Fischl, 2012), with manual corrections for stroke patients, and included registration to standard templates and mapping of cortical parcellations utilizing the Schäfer-Yeo (100 nodes) parcellation (Schaefer et al., 2018). DWI processing involved estimating fiber orientation distributions, and intensity normalization. Structural connectomes were created using (Tournier et al., 2019), incorporating tractography with iFOD2 algorithm and 3-tissue anatomically constrained tractography, followed by tract density image computation and Spherical deconvolution-informed filtering of tractograms (SIFT2 algorithm) (Smith et al., 2015). To define the connection weights between the cortical nodes in the structural connectivity matrices, we used the weighted streamline count, which represents the number of white matter streamlines connecting the nodes. To account for the potential effects of varying connection strengths, we applied a logarithmic transformation to the weighted streamline counts. This transformation reduces the impact of extreme values and scales the data to a more uniform range. We normalized the resulting values to ensure comparability across participants.

For gradients computation, we first constructed a group-level gradient component template by averaging the structural connectivity matrices across all participants and timepoints. This template served as a reference to ensure the alignment of individual gradient components aimed at facilitating the comparison of connectivity patterns among subjects. Affinity matrices for individual connectomes were formed using a normalized angle kernel. To align the individual gradient components to the group template, we used Procrustes alignment. Eigenvectors, determined via diffusion map embedding, projected connectome features into gradients. Gradient maps were computed at parcel level (Schaefer n=100). Accordingly, parcels with similar connectivity cluster together, while parcels with different connectivity locate farther apart in the gradient space. We analyzed separately intra- and inter-hemispheric cortical gradients. This approach allowed us to examine the differences in structural connectivity patterns within and between the two hemispheres. Intra-hemispheric gradients were computed within each hemisphere separately, while inter-hemispheric gradients were calculated across the hemispheres. Previous studies selected principal gradients within a range spanning from 2 to 4, aimed at capturing the most significant patterns in data (Larivière et al., 2024; Zarkali et al., 2021).

To ensure the gradients we derived align with established findings (Zarkali et al., 2021), we computed structural gradients in the healthy population, and we selected the top and bottom 10% of the average gradient values. These parcels were qualitatively compared with those commonly reported in previous literature.

**S1.5. Microstructural latent space computation**

DTI and NODDI maps (n=7) from the healthy controls (session 1) were first normalized to the MNI map. For this analysis we first registered each diffusion metric to the MNI space through the ANTs suite (Avants et al., 2008). Structural T1w images were first non-linearly registered to the MNI template and used as reference space for a rigid transformation of the b0 DWI map. The registration matrices were then used to normalize the diffusion maps in a single step with a 2mm resolution. For each metric from DTI and NODDI we computed an average map. The resulting control group-averaged maps were vectorized, resulting in a single matrix of n×m (n=number of maps, m=number of voxels). Voxel values of each map were z-scored according to its SD and mean values. Voxels with values higher than 5 SD were removed from the analysis to avoid potential bias in the computation of the factors. Before running the factorial analysis, we computed the Kaiser-Meyer-Olkin (KMO) test to assess the sampling adequacy of the PCA. Finally, the factorial analysis was run, and the first factors explaining more than 90% of variance were retained for subsequent analysis.

Stroke diffusion maps from both time points (acute and 3 months) and controls data from session 2 were normalized according to the same procedure described above and projected to the latent space computed in session 1 of the controls. This procedure ensure that each participant was projected in the same space, avoiding potential mismatch in the factorial space when computed at individual level. Further, spatial correlation between the projected maps and the template were computed to identify potential outliers. A cut-off of 0.2 was chosen to identify patients distant from the normative factorial maps.

Finally, to assess the stability of the latent structure reported in HC session 1, we performed the same analysis at the averaged-group level independently for diagnosis and timepoint (session 1 and session 2 from controls; acute and follow-up for stroke). Correlational analyses were performed to assess the similarity of the latent space

**S1.6. Human Connectome Project Diffusion Data**

HCP diffusion dataset was acquired through a 3T Magnetic Resonance Siemens Skyra “Connectome” scanner at Washington University in St. Louis. DWI images included a three shells diffusion scheme acquired via a whole-brain coverage; b-values=1000, 2000, and 3000 s/mm2; diffusion sampling directions of 90 directions per shell; in-plane resolution of 1.25 mm, with a slice thickness of 1.25 mm. T1-weighted images were acquired using a 3D MPRAGE sequence: time repetition (TR) = 2400 ms; time echo (TE) = 2.14 ms; voxel size = 0.7 mm isotropic. For detailed imaging parameters specific to the HCP data, please refer to (Van Essen et al., 2012). HCP data were preprocessed according to HCP core steps, including motion and eddy current correction, normalization, and skull striping. The preprocessing protocol and parameters as applied in the HCP can be found in Glasser et al., (2013).

The same microstructural approach applied to stroke patients was derived for HCP data. DTI and NODDI maps (n=7) were fed in the same latent factorial model to assess the reproducibility of this new space. For HCP, the latent diffusion factor was computed at individual level, thus resulting in 1064 latent maps. An averaged loading matrix was computed across the individual matrices and compared with the stroke cohort. Finally, the latent space was assessed using group-averaged diffusion maps in MNI space to assess the reproducibility at group-level.

**S1.7. Controls and stroke (acute) differences in gradients relationship**

For the structural equation modeling, data was prepared by creating an interaction term (G1 and Group) to capture potential moderating effects of group on the relationship between G1 and G2. This term was calculated as the product of the G1 value and a binary variable for group membership, where HC were coded as 0 and Stroke participants as 1. The SEM model included the following paths: a direct effect of G1 on G2, an effect of Group on G2, and the interaction effect, G1-group, on G2. Each network model was fit using the Sequential Least Squares Programming optimization method. Fit statistics were computed to evaluate model convergence and quality. For each network, we extracted parameter estimates, standard errors, z-values, and p-values for each pathway, allowing us to quantify the influence of group on the relationship between G1 and G2 and test the interaction hypothesis directly.

**S1.8. Whole brain comparison**

We compared WM latent diffusion factors at whole brain level between stroke patients (acute stage) and age-matched HC (from the WashU dataset). First, an “exclusion mask” was created based on the frequency map: lesioned voxels in more than 10% of the stroke cohort were excluded from further analysis (n=2 patients). Moreover, for each patient, factor maps were lesion masked. We employed a threshold-free cluster enhancement approach with n=1000 permutations and p<0.05 FWE for voxel-wise factorial analysis.

**S1.9. Sensitivity analysis: subcortical vs non-subcortical stroke lesions**

*S1.8.1 Diffusion map stability*

First, we assessed the reproducibility of the DTI and NODDI maps across the two sessions in the HC group from the WashU cohort. The distribution of spatial correlations among individuals was computed separately for FA, ODI, and ICVF maps in the WM and GM regions. Additionally, at the voxel-wise level, we computed the error maps (distances) between the two sessions for these DTI and NODDI maps. High correlations and low errors indicate high stability of the diffusion maps in the HC cohort over a 1-month interval.

*S1.8.2 Subcortical vs non-subcortical lesions*

We performed a sensitivity analysis comparing stroke at the acute stage and controls through an iterative process. Stroke patients were split into subcortical and non-subcortical groups (cortical or cerebellar lesions). Non-subcortical lesions were defined as lesions with no overlap with a subcortical mask from (Tian et al., 2020), while for the subcortical group we selected patients with an overlap above 20% of their lesion with the subcortical mask (the threshold was chosen to obtain an adequate sample size for performing the comparison). A one-way ANOVA with network and group factors, covarying for lesion volume, was performed to compare GD values between subcortical and non-subcortical groups. Further, an iteration approach was applied to assess the robustness of the results comparing stroke (acute) and controls. Specifically, at each iteration we removed a patient based on the ratio between lesions’ overlap with a subcortical mask (from Tian et al., 2021) and the lesion size (until reaching a stroke sample equal to the 15% of the original dataset). This analysis was repeated for both intra- and inter-connectivity structural gradients and at both network and parcel-levels comparing the subsample stroke iterative sample with the control cohort (kept fixed). The same analysis was applied at parcel-wise analysis, correcting p-values for FDR.

**S2. SUPPLEMENTARY RESULTS**

**S2.1. Structural connectivity gradients**

From an anatomical perspective, _intra_G1 described an axis spanning from positive loadings in the occipital lobe, through temporal cortex, to negative loadings in the frontal cortex. On the contrary, the frontal cortex represented the positive axis of _intra_G2 with the negative axis located in the parietal cortex. Finally, the positive epicenter of _intra_G3 was in the superior temporal gyrus, with the negative axis spanning to the occipital and medial frontal cortex (***Fig. 3*** *from the main text*). For the inter-hemispheric analysis, _inter_G1 was characterized by an axis ranging from the frontal lobe to the occipital and posterior parietal cortices. The second gradient (_Inter_G2) loads mainly in the medial regions, including the central sulcus area, and negative values in both anterior and posterior peripheral regions, suggesting a delineation of primary sensory/motor areas from associative areas. Finally, _inter_G3, shows an extreme of the axis located in the ventral parts of the brain, particularly the inferior frontal and temporal lobes, and the opposite in dorsal areas, including the superior frontal and parietal regions, illustrating a ventral-dorsal brain axis (***Fig. 3*** *from the main text*).

**S2.2. Support vector machine assessing gradient space in controls vs acute stroke**

*S2.2.1. Intra-hemispheric connectivity structural gradients*

The best-performing parameters were identified as C=5.7, gamma: auto, and kernel: rbf, achieving an LOOCV accuracy of 83% and a test accuracy of 89%. When applied to the entire dataset, the model demonstrated a classification accuracy of 92%, confirming its robustness.

*S2.2.2. Inter-hemispheric connectivity structural gradients*

The best-performing parameters were identified as C=0.7, gamma: auto, and kernel: poly, achieving an LOOCV accuracy of 93% and a test accuracy of 96%. When applied to the entire dataset, the model demonstrated a classification accuracy of 97%, confirming its robustness.

**S.2.3. Acute stroke vs controls in gradient organization**

*Intra-connectivity network structural organization*

We compared absolute network-gradient values between HC and stroke. Significant _intra_G1 diagnosis and network effects were reported for both ipsilateral (group: F=37.9; p<0.001; network: F=102.9; p<0.001) and contralateral (group: F=13.1; p<0.001; network: F=107.2; p<0.001) hemispheres to the lesion. No significant diagnosis × network effect was observed for all the analysis (p>0.05), suggesting widespread effects across networks. _intra_G2 showed similar results for both ipsi- (diagnosis: F=56.6, p<0.001; network: F=13.0, p<0.001) and contra-lesional hemisphere (diagnosis: F=9.32, p=0.002; network: F=34.4, p<0.001). A significant diagnosis × network effect was observed for the ipsilesional gradient (F=2.49; p=0.022), where all networks were significantly different, except the visual network (p<0.004 for all the comparisons, Bonferroni corrected). This is explained by the mainly central location of the average group lesions that relatively spared the posterior regions of the brain. For the _intra_G3 we found similar patterns, with significant diagnosis and network effects for both the ipsi- (diagnosis: F=33.1, p<0.001; network: F=6.82, p<0.001) and contralesional hemisphere (diagnosis: F=24.4, p<0.001; network: F=16.9, p<0.001). No significant diagnosis × network was reported (p>0.05). All these results were adjusted for age, gender and education. Overall, these results suggest a whole brain widespread difference of structural gradients between stroke and age-matched controls.

The effect of diagnosis echoed in the ipsilesional hemisphere when comparing GD values between controls and stroke (F=65.64; p<0.001), while no network (p=0.07) and diagnosis × network (p>0.05) effects were reported (p>0.05). For the contralesional hemisphere a significant effect was reported for both diagnosis (F=28.8; p<0.001) and network (F=3.72; p=0.001) but not for the interaction term (p>0.05). Results are reported in **Supplementary Fig. S4**.

*Inter-connectivity network structural organization*

For both ipsi and contralesional hemispheres to the lesion, _inter_G1 showed significant group (ipsi: F=6.21; p=0.013; contra: F=12.4; p<0.001), and network effects (F=77.8; p<0.0001), with no interactions (p>0.05). Similarly, _inter_G2 showed significant group (ipsi: F=39.3; p<0.001 contra: F=44.5; p<0.001), and network (ipsi: F=33.5; p<0.001; contra: F=28.2; p<0.001) effects while no interactions were reported (p<0.05). For _inter_G3 we reported a significant group (ipsi: F=24.7; p<0.001; ipsi: F=44.0; p<0.001), and network (ipsi: F=13.3; p<0.001; contra: F=16.0; p<0.001) effects, with no significant interactions (p<0.05).

As for the intra-hemispheric results, analysis on GD values were in line with these results, showing a significant effect of diagnosis (F=54.5; p<0.001). The effect of diagnosis was echoed when comparing GD values between controls and stroke (ipsi: F=32.7; p<0.001; contra: F=54.5; p<0.001), while no network (p>0.05) and diagnosis × network effects (p>0.05) were reported (**Supplementary Fig. S4**).

**S2.4. Microstructural latent space in the HCP dataset**

In the HCP cohort we consistently reported the 3 main latent diffusion factors observed in the WashU cohort. Specifically, the first factor (dwiF1) explained 47% of the variance loading of diffusion DTI/NODDI outcomes; the second factor (dwiF2) - 32% of the variance explained - loaded on FA and ODI; the third factor (dwiF3) loaded mainly on FA and ICVF, with a strong contribution of ISOVF (in contrast with WashU results), explain around 15% of the variance. This pattern was consistently reported both at group level (***Fig. 4*** *from the main text*) and single subjects’ space (**Supplementary Fig S3**). For less than 5% of the sample, factors computed in the native space showed low spatial similarity with the group-averaged factors, indicating a robust stability also at single subject level (**Supplementary Fig S3**).

**2.5. Voxel-wise microstructural differences between acute stroke and controls**

A significant difference was reported for stroke (acute) patients compared to controls for the second latent diffusion component (_dwi_F2). Compared with the Johns Hopkins University (JHU)-ICBM-DT Atlas, significant regions mapped on the forceps major and minor, and the bilateral superior and inferior longitudinal fasciculi. The location of these major white matter tracts that connect the two hemispheres (forceps) and anterior to posterior regions (longitudinal fasciculi) is consistent with the average central location of the lesions deep in the white matter and their widespread effect on intra- and inter-hemispheric structural gradients. Results are shown in **Supplementary Fig. S6.**

**2.6. Structural gradients sensitivity analysis: subcortical vs non-subcortical**

Since stroke lesions occur predominantly in WM it is possible that changes in structural gradients were sensitive to lesion location. Lesions were automatically classified as subcortical or non-subcortical (according to the overlap with a prespecified mask). Stroke-related changes in cortical gradients were re-analyzed by systematically removing more cortical or more subcortical lesions through an iterative process. The ANOVA, executed by iteratively excluding a patient with the lowest subcortical involvement (expressed as the ratio between lesion voxels mapping to the subcortical mask and lesion size), demonstrated a consistently significant group effect (stroke acute vs HC) for all intra-connectivity gradients considered. This remained consistent across iterations, irrespective of the lesion side, thus suggesting an independent effect of lesion topology. Similarly, all gradients exhibited a significant group effect for inter-connectivity patterns involving the ipsilateral hemisphere, and _intra_G2 and _intra_G3 for the contralateral side, thus corroborating the findings from the entire cohort. The results are presented in **Supplementary Fig. S6**. Similarly, gradient values were similar between patients with lesions mapping in the cortical or subcortical region (**Supplementary Fig. S6).**

**Table S1. Linear Mixed Model results for the ITT-L sample (GD acute vs follow-up)**

|  | **Estimate** | **95% CI** | **SE** | **T-stat** | **p-value** |
| --- | --- | --- | --- | --- | --- |
| Ipsilateral connectivity | | | | | |
| _intra_GD | *0.024* | *0.010 – 0.037* | *0.007* | *3.406* | *0.001* |
| _inter_GD | *0.012* | *0.004 – 0.020* | *0.004* | *3.035* | *0.003* |
| Contralateral connectivity | | | | | |
| _intra_GD | *0.018* | *0.008 – 0.028* | *0.005* | *3.431* | *0.001* |
| _inter_GD | *0.014* | *0.004 – 0.025* | *0.005* | *2.667* | *0.008* |
| Microstructural latent factor dwiF1 | | | | | |
| SDC 40% | 0.004 | -0.029 – 0.037 | 0.017 | 0.259 | 0.798 |
| SDC 60% | 0.001 | -0.038– 0.040 | 0.020 | 0.062 | 0.951 |
| SDC 80% | -0.005 | -0056 – 0.045 | 0.026 | -0.200 | 0.843 |
| Microstructural latent factor dwiF2 | | | | | |
| SDC 40% | -0.033 | -0.100 – 0.034 | 0.034 | -0.957 | 0.348 |
| SDC 60% | -0.024 | -0.102 – 0.053 | 0.040 | -0.615 | 0.544 |
| SDC 80% | -0.014 | -0.116 – 0.088 | 0.052 | -0.277 | 0.748 |
| Microstructural latent factor dwiF3 | | | | | |
| SDC 40% | *0.060* | *0.005 – 0.115* | *0.028* | *2.124* | *0.043* |
| SDC 60% | *0.077* | *0.022 – 0.133* | *0.028* | *2.747* | *0.011* |
| SDC 80% | *0.078* | *0.006 – 0.150* | *0.037* | *2.128* | *0.043* |

**Table S2. Linear Mixed Model results for the PPA-L sample (GD acute vs follow-up)**

|  | **Estimate** | **95% CI** | **SE** | **T-stat** | **p-value** |
| --- | --- | --- | --- | --- | --- |
| Ipsilateral connectivity | | | | | |
| _intra_GD | *0.023* | *0.008 – 0.034* | *0.007* | *3.057* | *0.002* |
| _inter_GD | *0.013* | *0.005 – 0.021* | *0.004* | *3.175* | *0.002* |
| Contralateral connectivity | | | | | |
| _intra_GD | *0.019* | *0.008 – 0.031* | *0.006* | *3.332* | *0.001* |
| _inter_GD | *0.015* | *0.003 – 0.026* | *0.006* | *2.529* | *0.012* |
| Microstructural latent factor dwiF1 | | | | | |
| SDC 40% | 0.007 | -0.028 – 0.041 | 0.018 | 0.392 | 0.699 |
| SDC 60% | 0.003 | -0.038 – 0.044 | 0.021 | 0.138 | 0.892 |
| SDC 80% | -0.006 | -0.061 – 0.048 | 0.028 | -0.229 | 0.821 |
| Microstructural latent factor dwiF2 | | | | | |
| SDC 40% | -0.030 | -0.100 – 0.040 | 0.036 | -0.835 | 0.413 |
| SDC 60% | -0.022 | -0.104 – 0.060 | 0.042 | -0.526 | 0.605 |
| SDC 80% | -0.009 | -0.117 – 0.099 | 0.055 | -0.163 | 0.872 |
| Microstructural latent factor dwiF3 | | | | | |
| SDC 40% | *0.060* | *0.002 – 0.118* | *0.03* | *2.026* | *0.056* |
| *SDC 60%* | *0.078* | *0.020 – 0.136* | *0.030* | *2.263* | *0.016* |
| SDC 80% | *0.076* | *-0.002 – 0.154* | *0.040* | *1.915* | *0.070* |

**Table S3. Linear regression analysis for the relationship between microstructural outcomes within the disconnected tracts and gradient divergent values.**

|  | **Estimate** | **SE** | **T-stat** | **p-value** | **95% CI** |
| --- | --- | --- | --- | --- | --- |
| Intra-hemispheric / Contralesional | | | | | |
| dwiF2 | 0.0787 | 0.036 | 2.203 | *0.034* | 0.006 / 0.151 |
| dwiF3 | -0.1471 | 0.054 | -2.721 | *0.010* | -0.257 / -0.038 |
| Intra-hemispheric / Ipsilesional | | | | | |
| dwiF2 | 0.1317 | 0.061 | 2.151 | *0.038* | 0.008 / 0.256 |
| dwiF3 | -0.2963 | 0.093 | -3.198 | *0.003* | -0.484 / -0.109 |
| Inter-hemispheric / Contralesional | | | | | |
| dwiF2 | 0.0389 | 0.040 | 0.979 | 0.334 | -0.042 / 0.120 |
| dwiF3 | -0.1897 | 0.060 | -3.144 | *0.003* | -0.312 / -0.067 |
| Inter-hemispheric / Ipsilesional | | | | | |
| dwiF2 | 0.0197 | 0.044 | 0.445 | 0.659 | -0.070 / 0.110 |
| dwiF3 | -0.1674 | 0.067 | -2.486 | *0.018* | -0.304 / -0.031 |

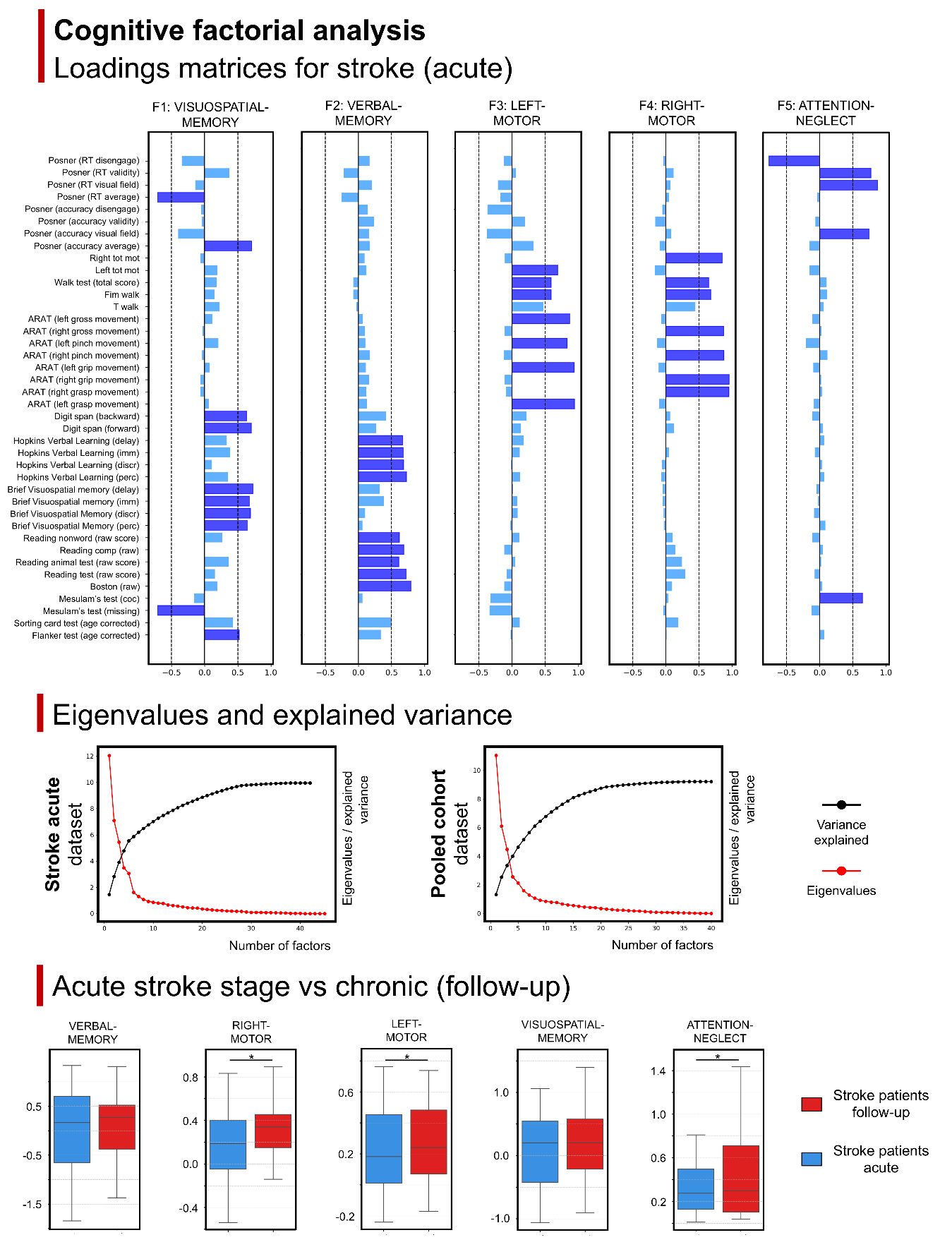

**Figure S1. Cognitive factors in the stroke sample**

Top panel: latent factors analysis limited to the stroke (acute) cohort. Center panels: Eigenvalues and variance explained for the stroke acute cohort (left) and the whole dataset (right). Bottom panels: Statistical comparison between stroke patients in acute and follow-up stages. * marks significant differences

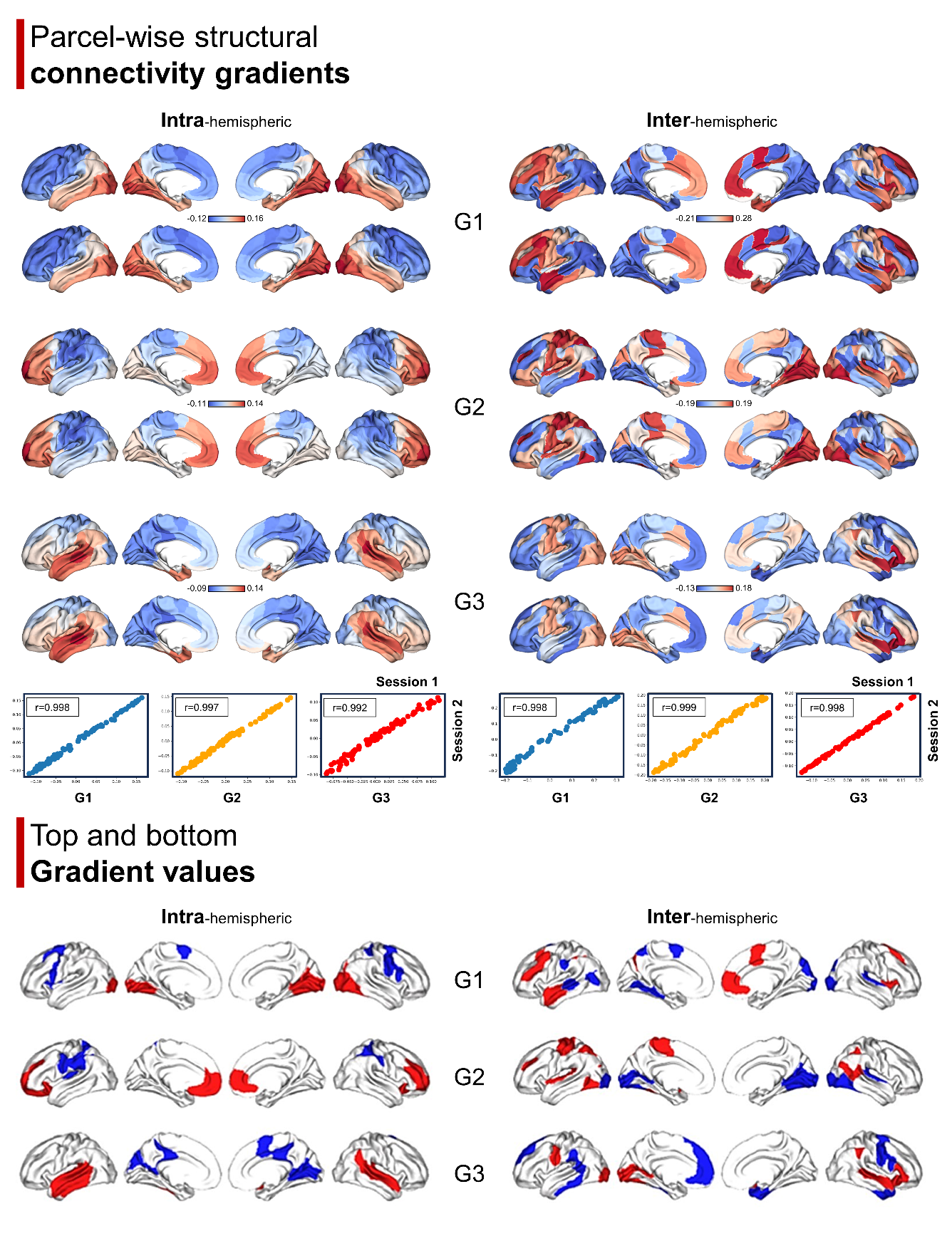

**Figure S2. Structural connectivity reproducibility and epicenters**

**Top panel**: intra- (left) and inter- (right) hemispheric structural connectivity gradients were computed independently for the healthy control sample during two visits. Averaged gradient templates for the timepoints are reported (first row: visit 1; second row: visit 2). The scatterplot illustrates the relationship between the two maps computed on the cohort at different visits. **Bottom panel**: top (red) and bottom (blue) 10% parcels from the structural connectivity gradients in the control sample for the intra- (left) and inter- (right) hemispheric gradients.

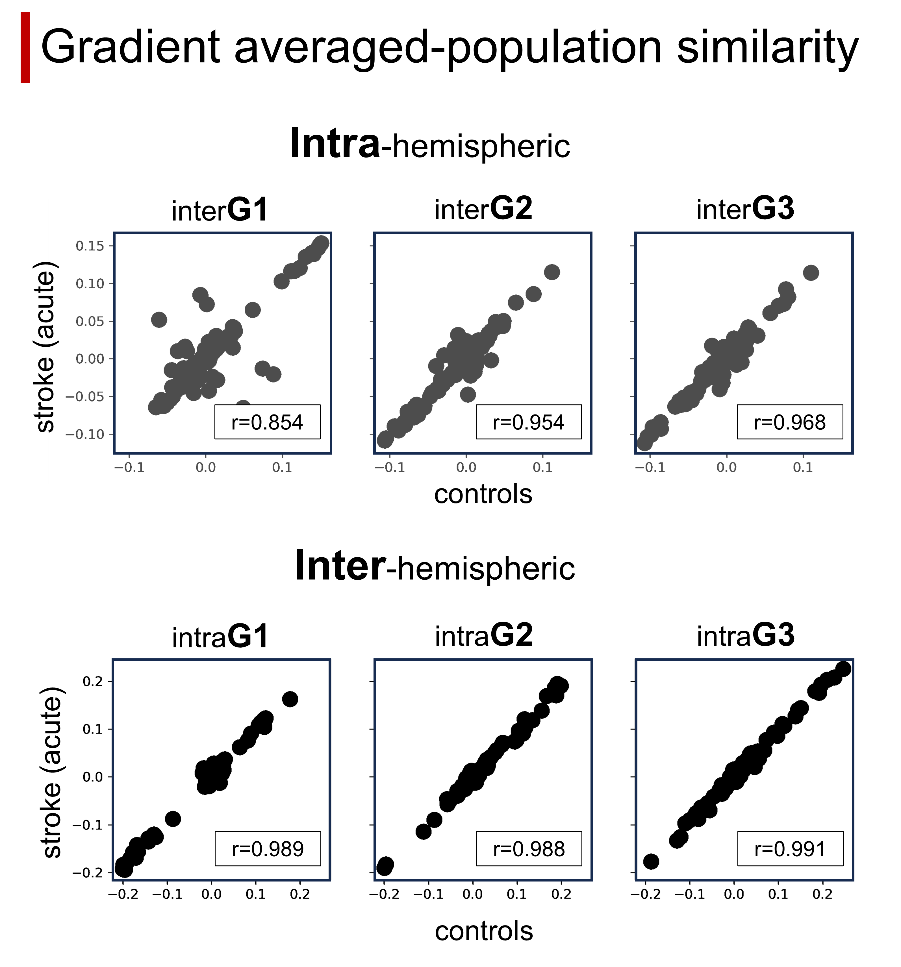

**Figure S3. Group average gradient-based similarity between stroke (acute stage) and controls.**

Intra-hemispheric (top row) and inter-hemispheric (bottom row) correlations across the three main structural gradient-averaged templates are reported. Each dot represents a parcel value from the controls group’s template (x-axis) and stroke population’s template (y-axis). Box plot reports the correlation coefficients (r).

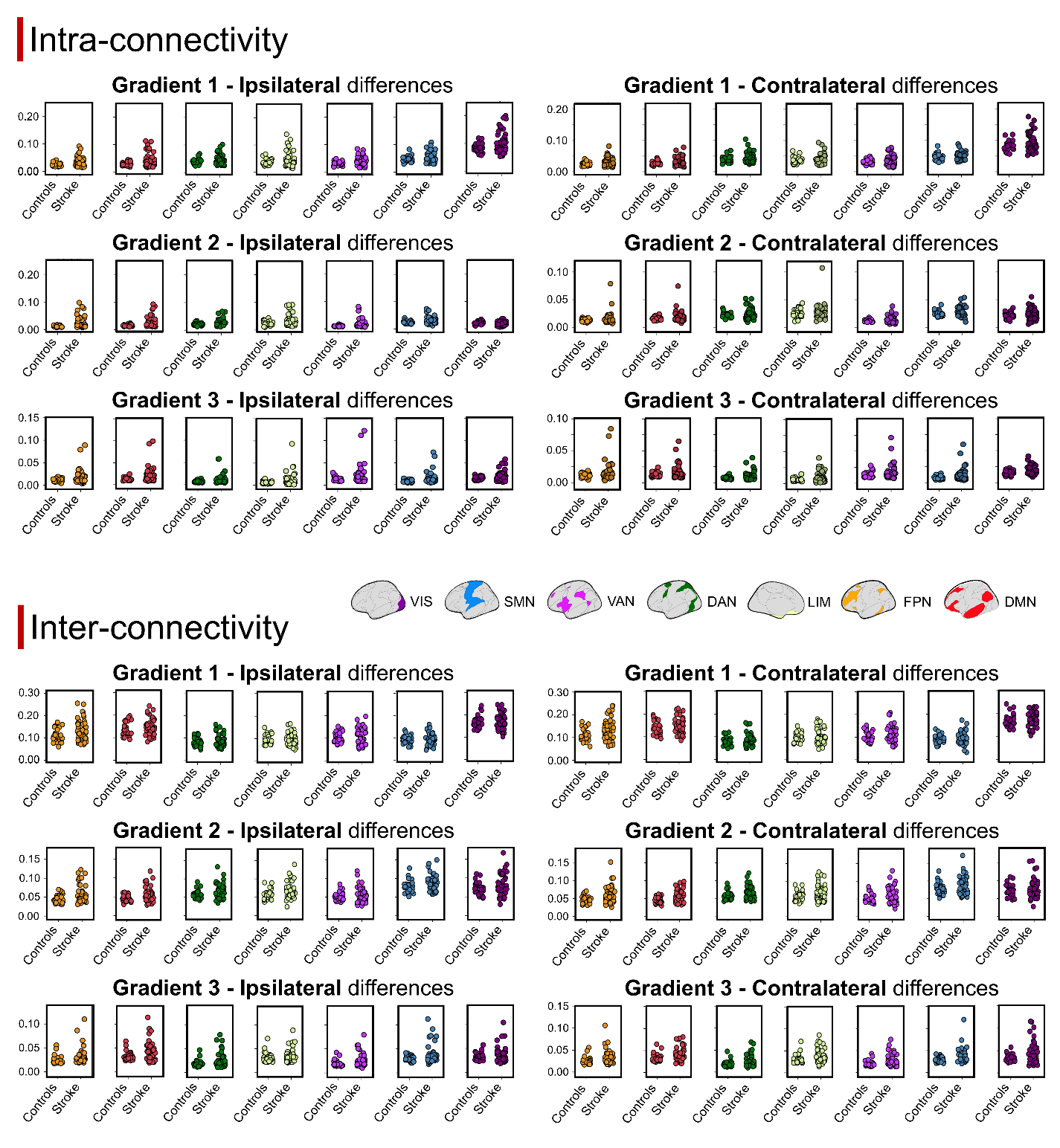

**Figure S4. Cross-sectional comparison between stroke (acute) and healthy controls.**

Top panel: intra-connectivity network-wise gradient values (absolute) comparison between the different groups (controls and stroke). Each scatterplot reports the averaged-network value for the individuals included in the comparison. Higher values mean higher distance from the canonical template. Bottom panel: The same analysis was repeated for the inter-connectivity structural gradients. Abbreviations: DMN - default mode network; DAN - dorsal-attention network; SMN - sensorimotor network; LMB - limbic network; VIS - visual network; FPN - frontoparietal network; VAN - ventral-attention. The network color code of the plots matches that of the projected parcels on the brain surface (middle panel).

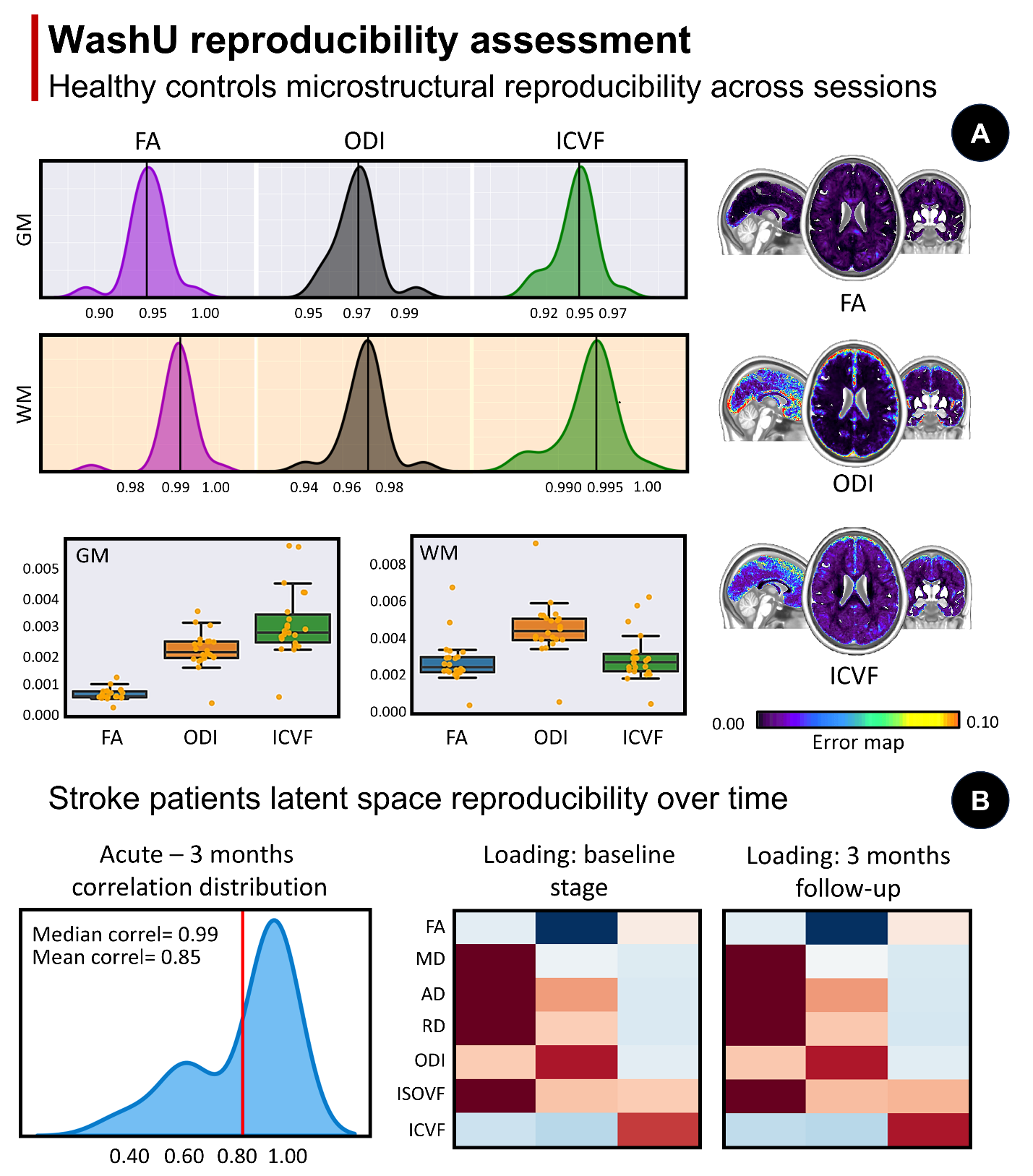

**Figure S5. Spatial and reproducibility properties of the diffusion latent space**

**Panel A**: spatial correspondence and error maps from the main diffusion maps computed from DTI and NODDI models (FA, ODI, ICVF). The distribution plot shows the spatial relationship between these maps computed in the first and second session from the WashU controls cohort. In the boxplot the error between these maps (distance) is depicted, along with the projection of error scores at voxel-wise level. **Panel B**: loadings of the latent factor space computed in stroke patients independently at the acute and follow-up stage (group-level factors). The distribution panel shows the correlation of the loadings extracted at acute and 3 months follow-up.

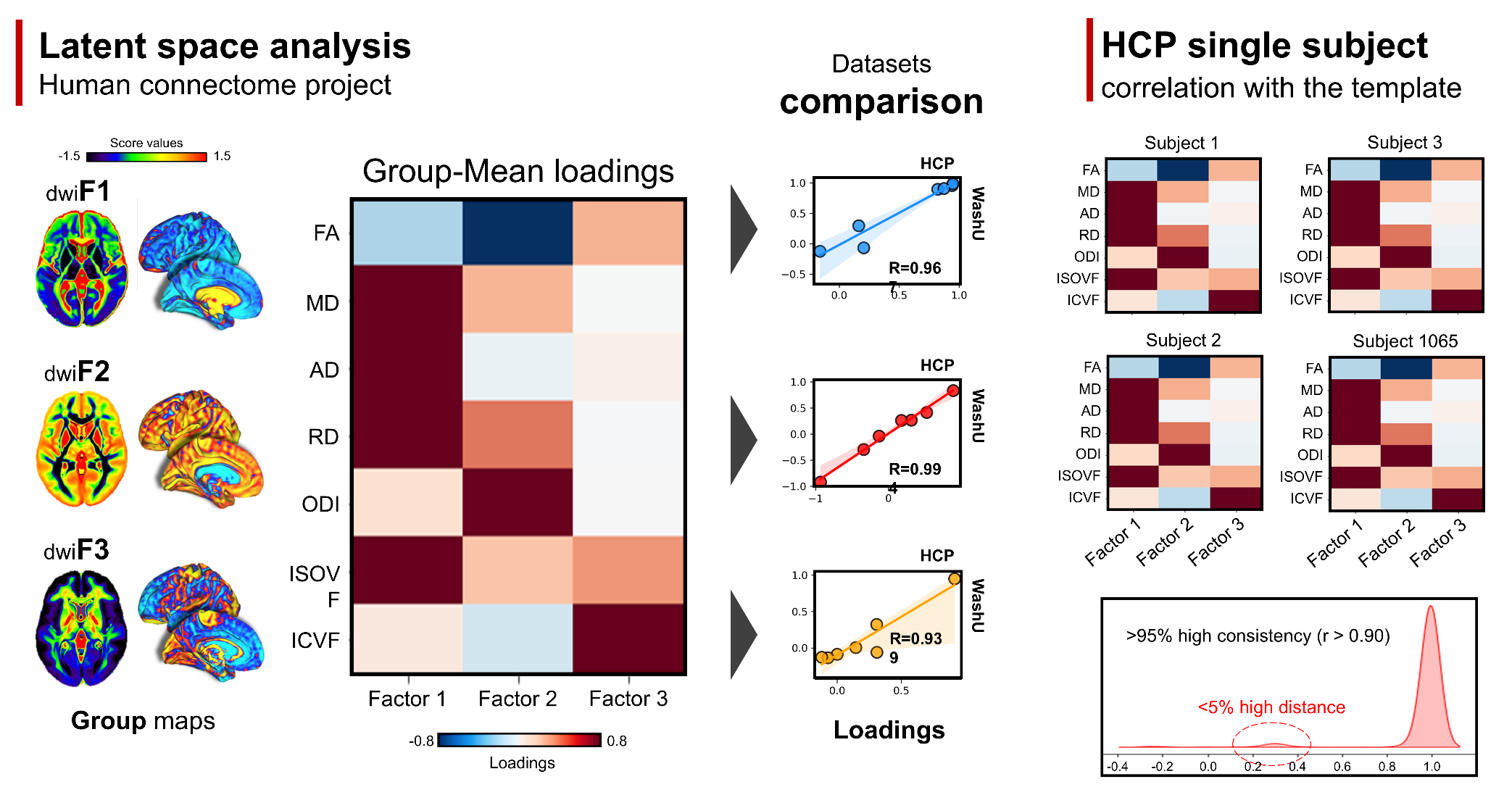

**Figure S6. Microstructural latent space diffusion in the Human Connectome Project**

**Left panels**: replication of microstructural factors in the Human Connectome Project dataset (n=1065). The single matrix depicts the loadings computed at group-average level. The weights are projected in volumetric and surface space for visualization purposes. **Right panels**: reproducibility of the latent space at individual level. Four different exemplative individuals from the HCP cohort are depicted (loadings). The correlation between individual loadings with the averaged-group template is reported in the distribution plot, showing that more than 95% of the sample showed a highly reproducible (r>0.95) latent space.

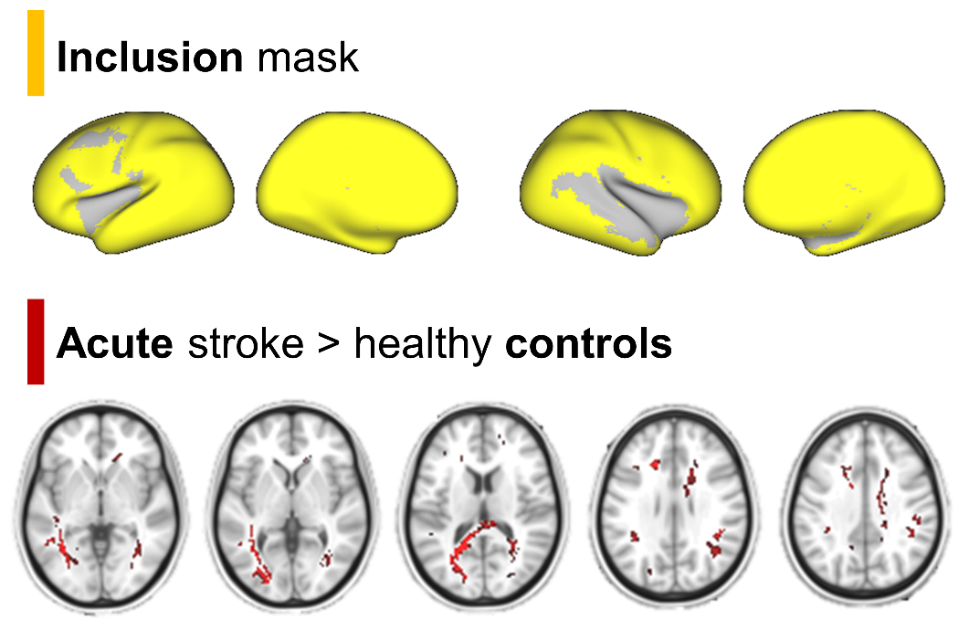

**Figure Supplementary S7. Voxel-wise differences between controls and stroke (acute).**

Top panel: an inclusion mask for was computed excluding lesioned voxels in more than 10% of the stroke sample (yellow highlights regions included in the mask). Bottom panels: voxel-wise (white matter) results comparing stroke and controls. Results were corrected at p<0.05 with multiple comparisons (FWE).

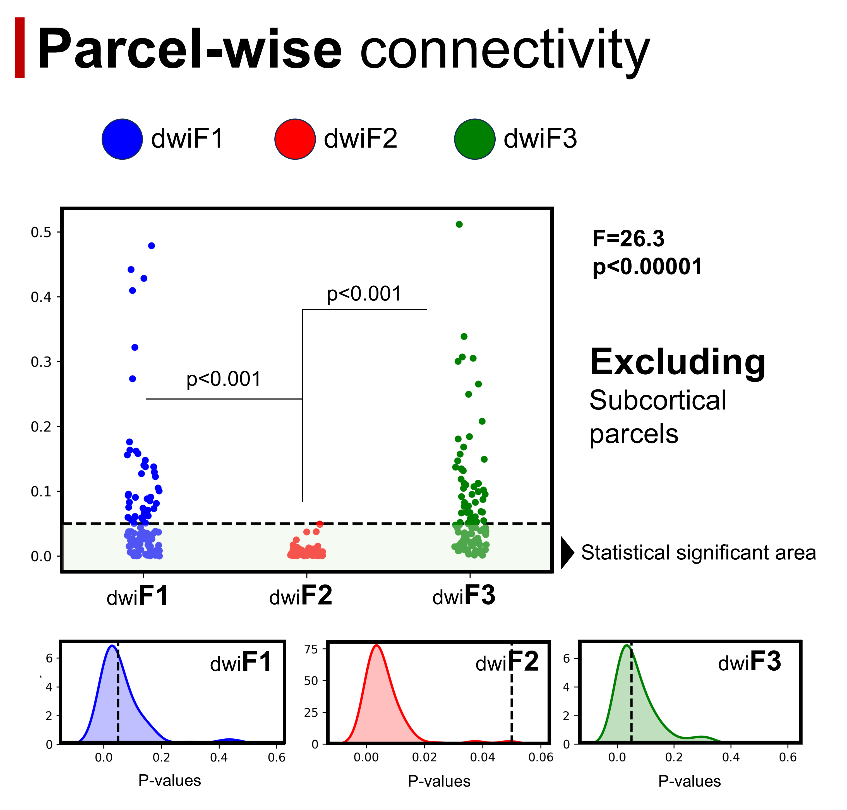

**Figure Supplementary S8. Disconnected vs non-disconnected parcels microstructural differences.**

Microstructural latent properties within parcels classified as disconnected *versus* not disconnected from the lesion. The procedure was iteratively repeated by randomly selecting the least disconnected parcel, excluding the subcortical parcels. Each plot in the panel shows the p-value associated with each iteration. Significant p-values are located in the bottom region (green), while non-significant p-values are in the top (red) zone. The KDE plot illustrates the distribution of p-values for each microstructural latent factor.

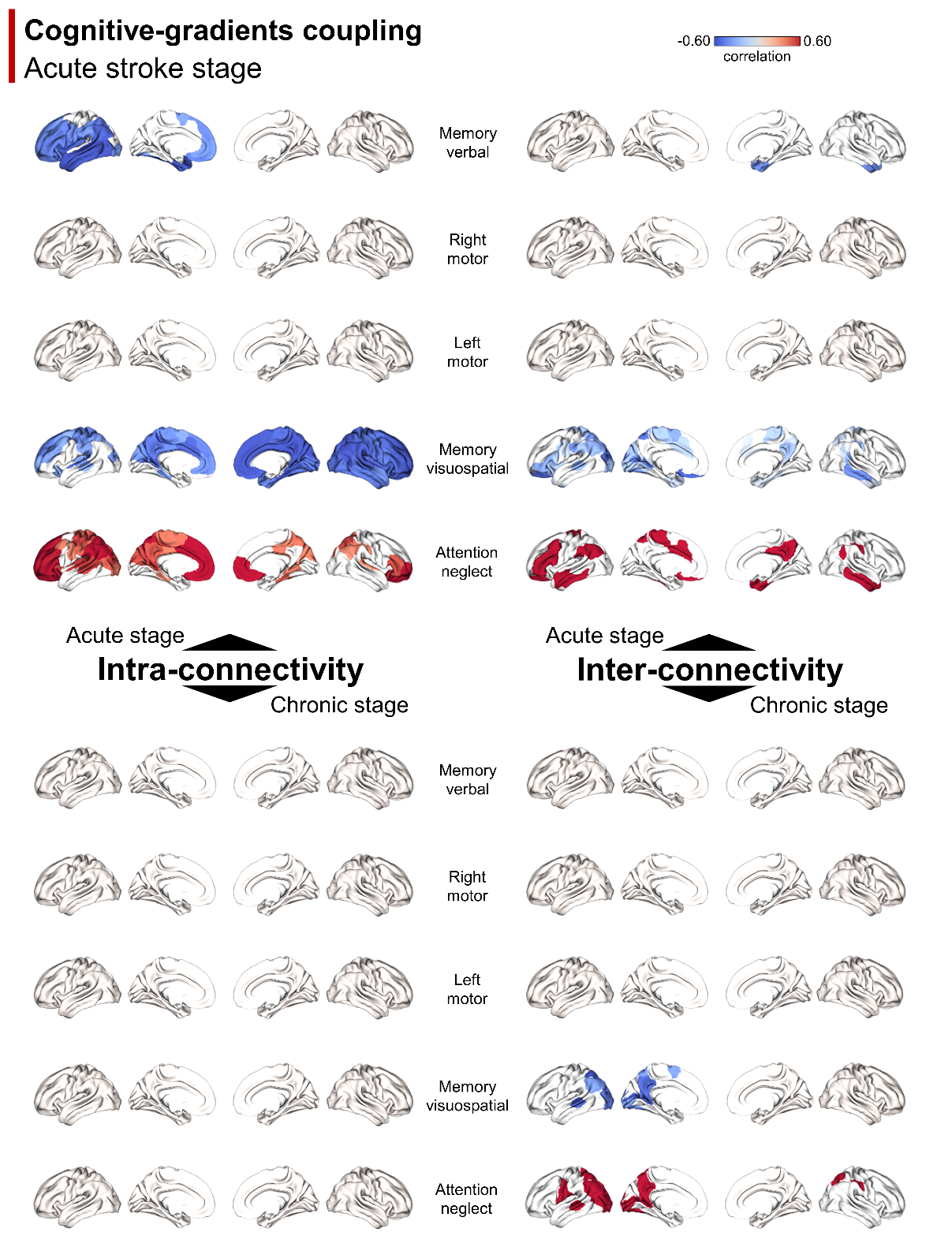

**Figure Supplementary S9. Parcel-wise cognitive-gradient relation for acute and chronic stages.**

The linear correlation between behavioral scores and GD values at the parcel level was computed for each behavioral factor. The figure show significant parcel-wise r-values (red: positive; blue: negative) surviving p<0.05 FDR. Results are reported for both intra- (left panels) and inter- (right panels) hemispheric connectivity gradients and for acute (top panels) and chronic (bottom panels) gradients.

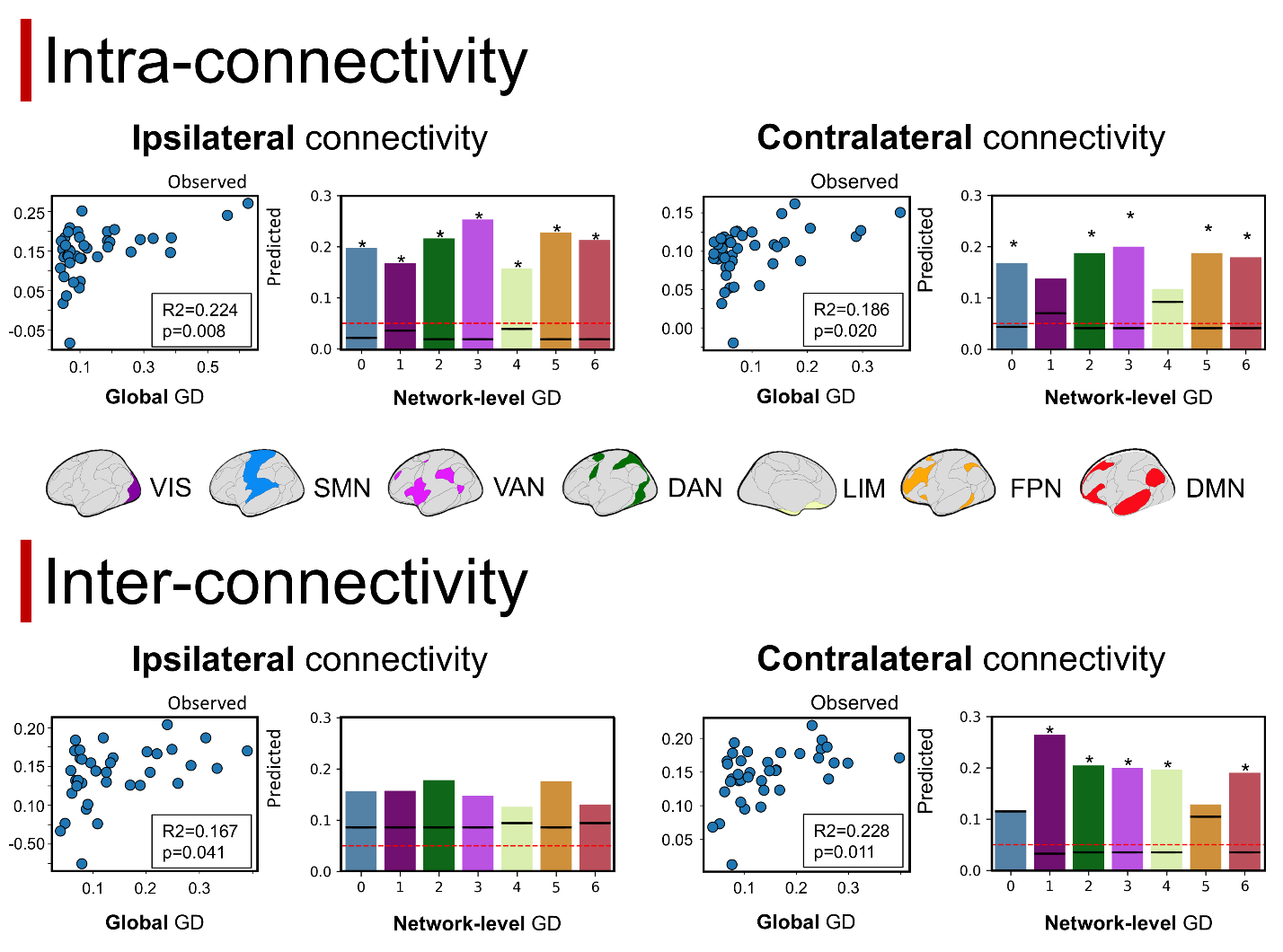

**Figure Supplementary S10. Structural gradients and microstructural relation in (acute) stroke.**

For each analysis considered, in the scatterplot, each point corresponds to the global GD value predicted (y-axis) by microstructural values within the disconnection mask and the observed value (x-axis). The same analysis was replicated at the network level (GD averaged across parcels belonging to the same network according to Yeo et al., 2011). For each model, the R² value is represented by a bar, and the associated p-value (FDR-corrected) is shown as a black line overlaid on each bar. The red dashed line indicates the significance threshold. An asterisk (*) marks significant models. The color of each bar matches the color of the networks represented on the surface (central panel).

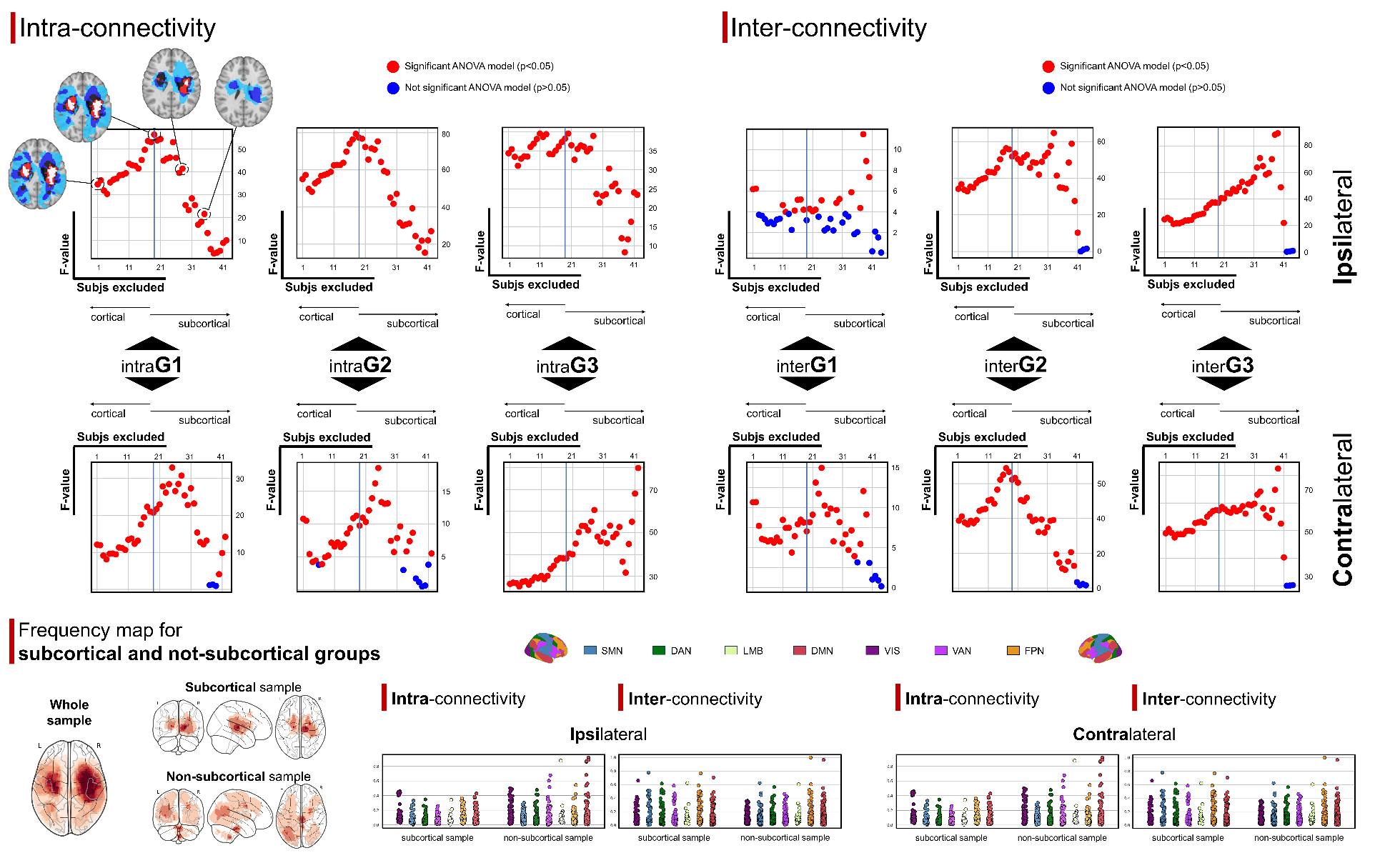

**Figure S11. Gradients differences at acute stage in subcortical and non-subcortical stroke.**

**Top panel**: stroke (acute stage) and controls were compared through an iterative process removing from the analysis the patient with the lowest subcortical lesion involvement. F-value from the group effects were reported for each iteration, showing a stable significant pattern across the dataset (red points: significant results (p<0.05); blue points: not significant (p>0.05);. Exemplative frequency maps from the resulting sample for specific iteration are reported and superimposed onto a standard brain template. **Bottom panel**: Frequency maps for the stroke sub-cohorts are reported in the left. No significant network-wise differences were reported for GD comparing subcortical and non-subcortical cohorts for both intra- (left) and inter- (right) connectivity gradients of the ipsi- (top) and contra- (bottom) lesion hemispheres (right panel).
