## Supplementary figures and images for "Longitudinal degeneration of microstructural and structural connectivity patterns following stroke"

### Figure 1

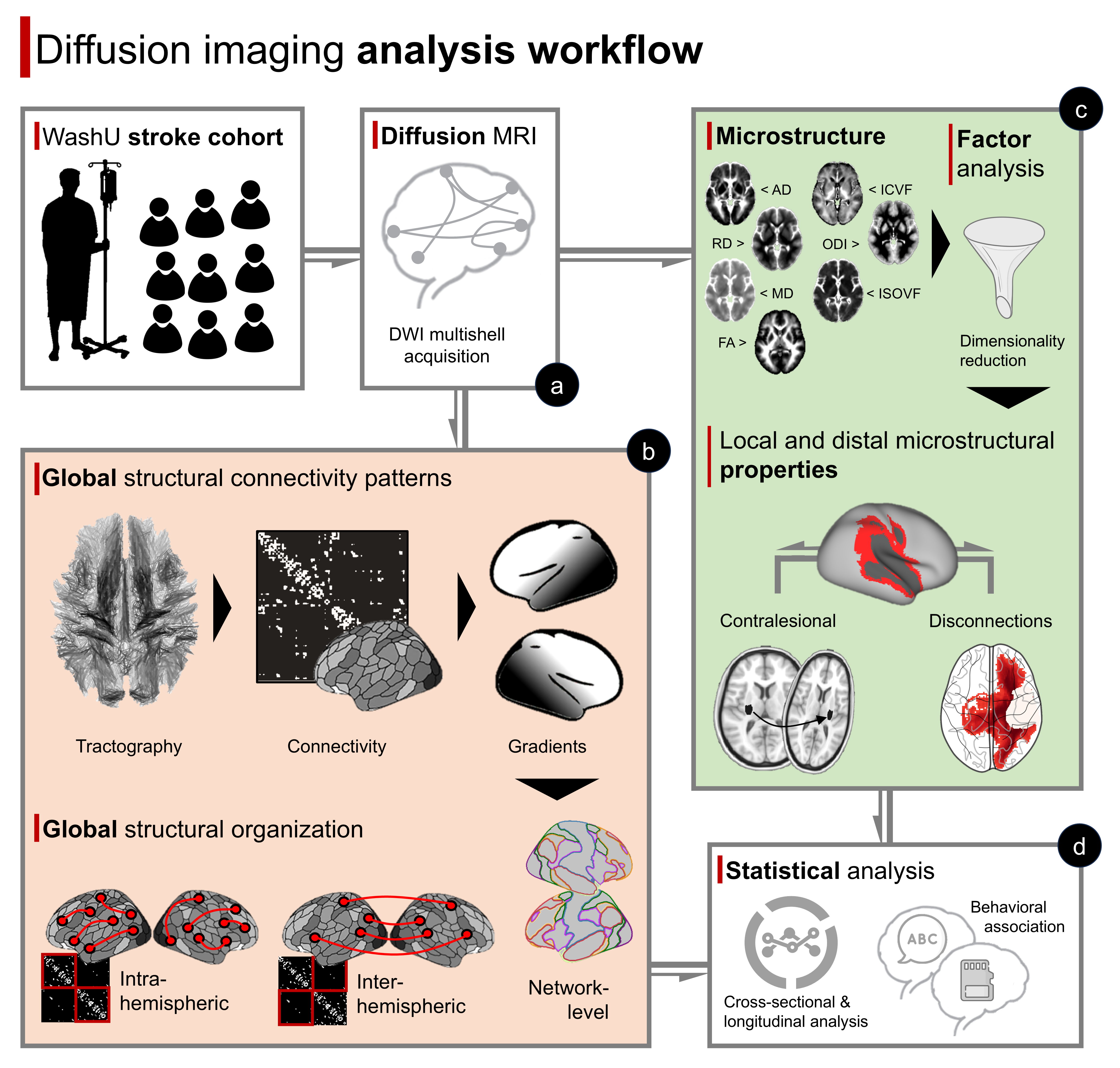

### Figure 2

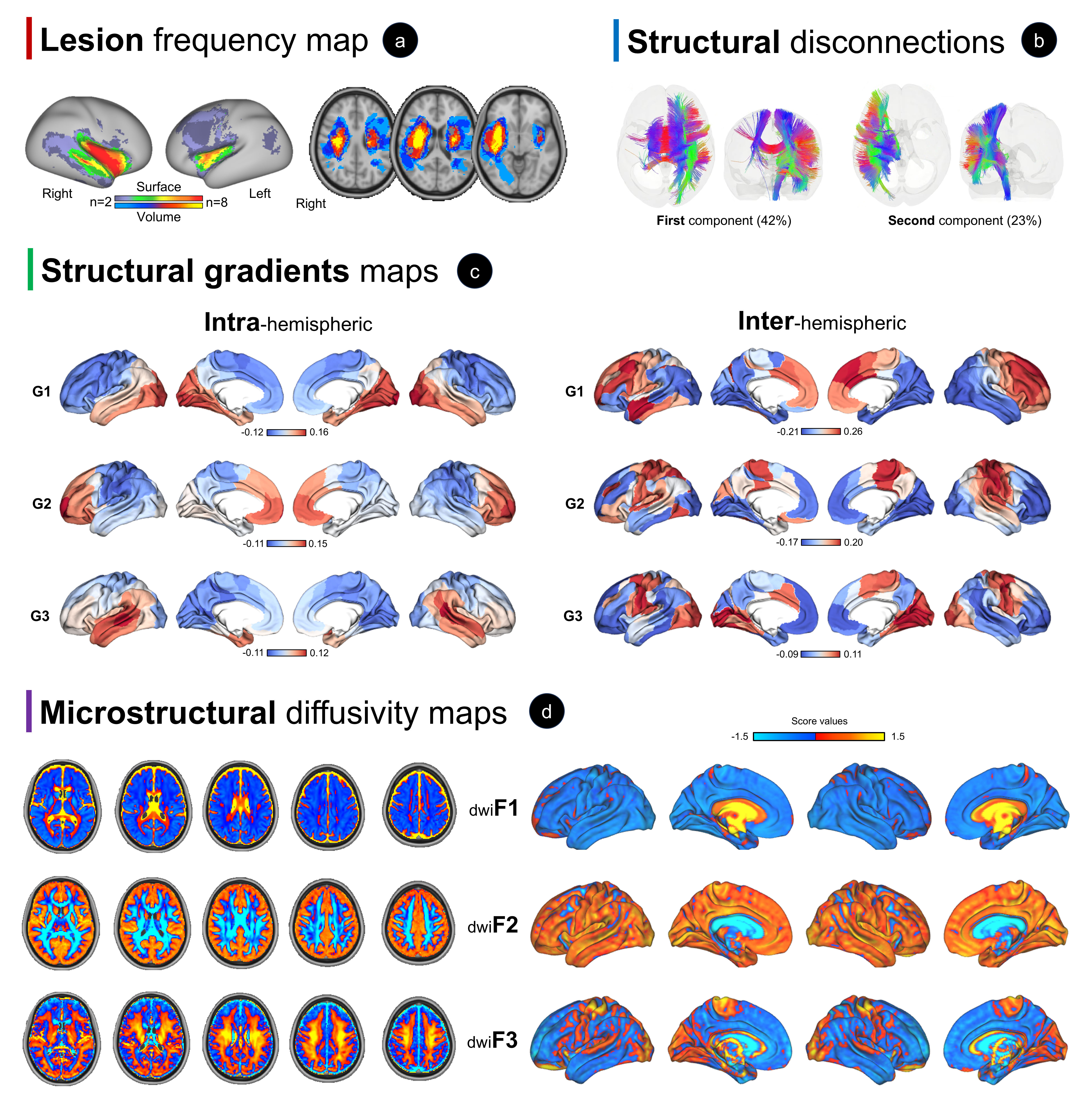

### Figure 3

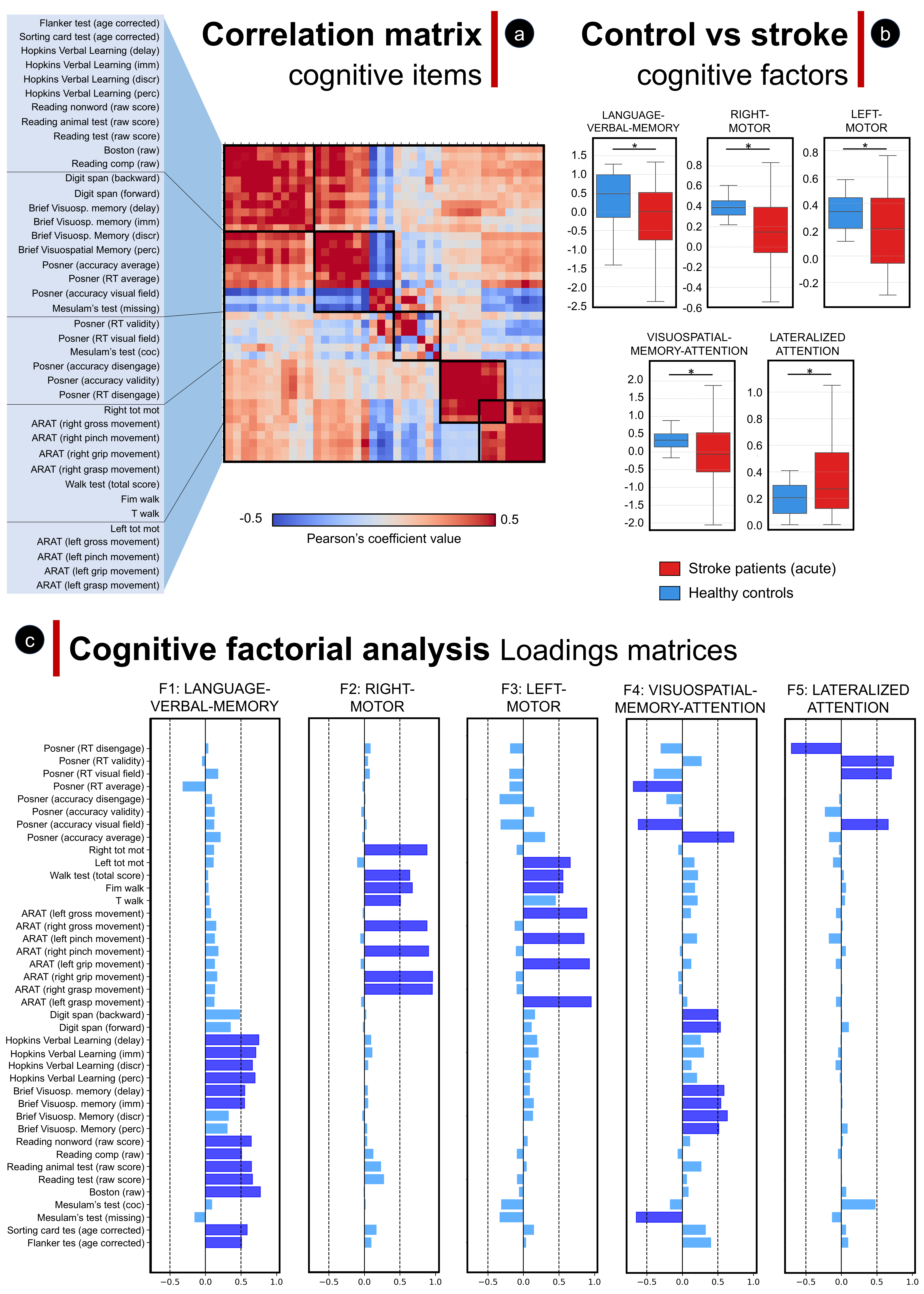

### Figure 4

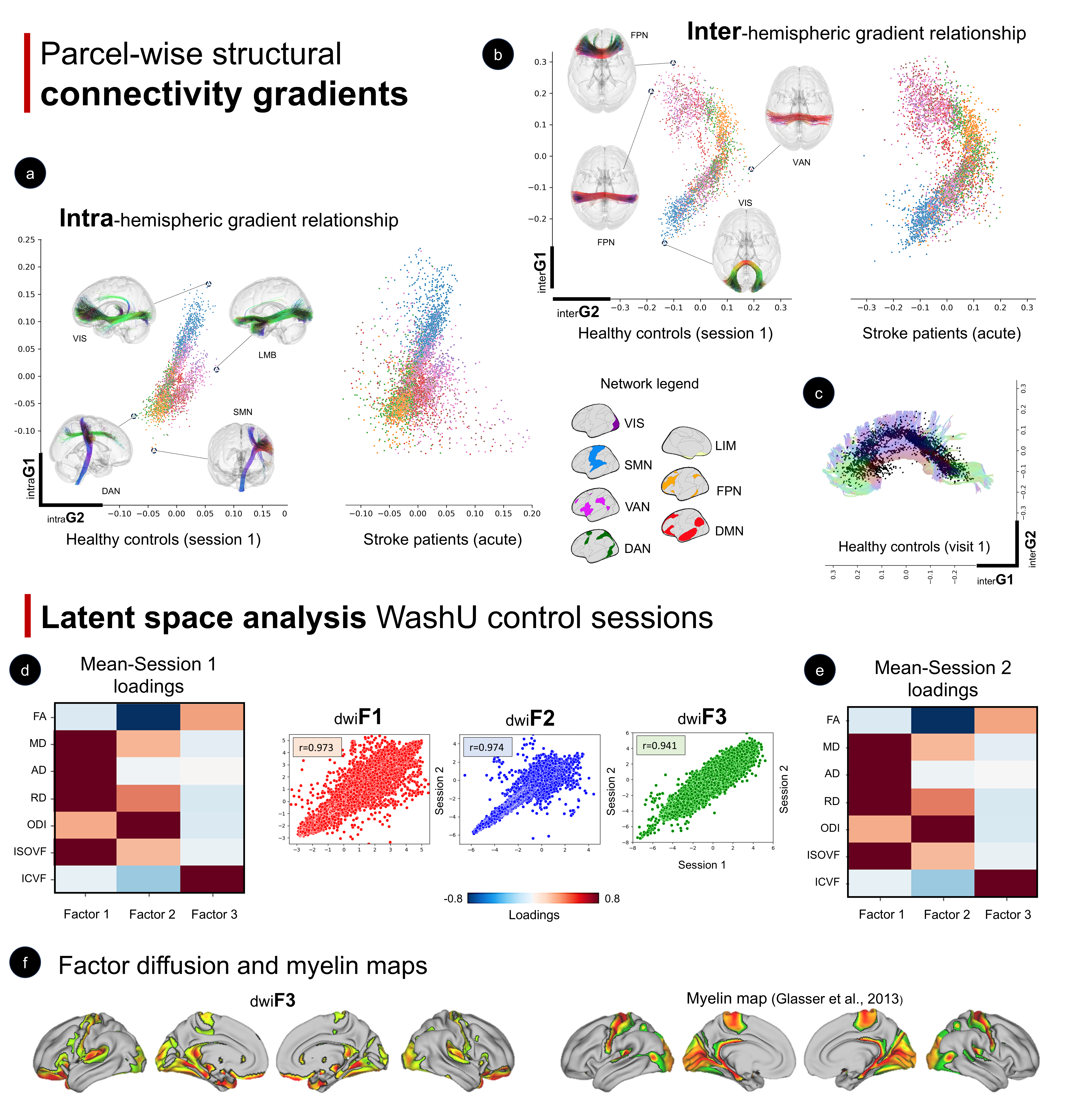

### Figure 5

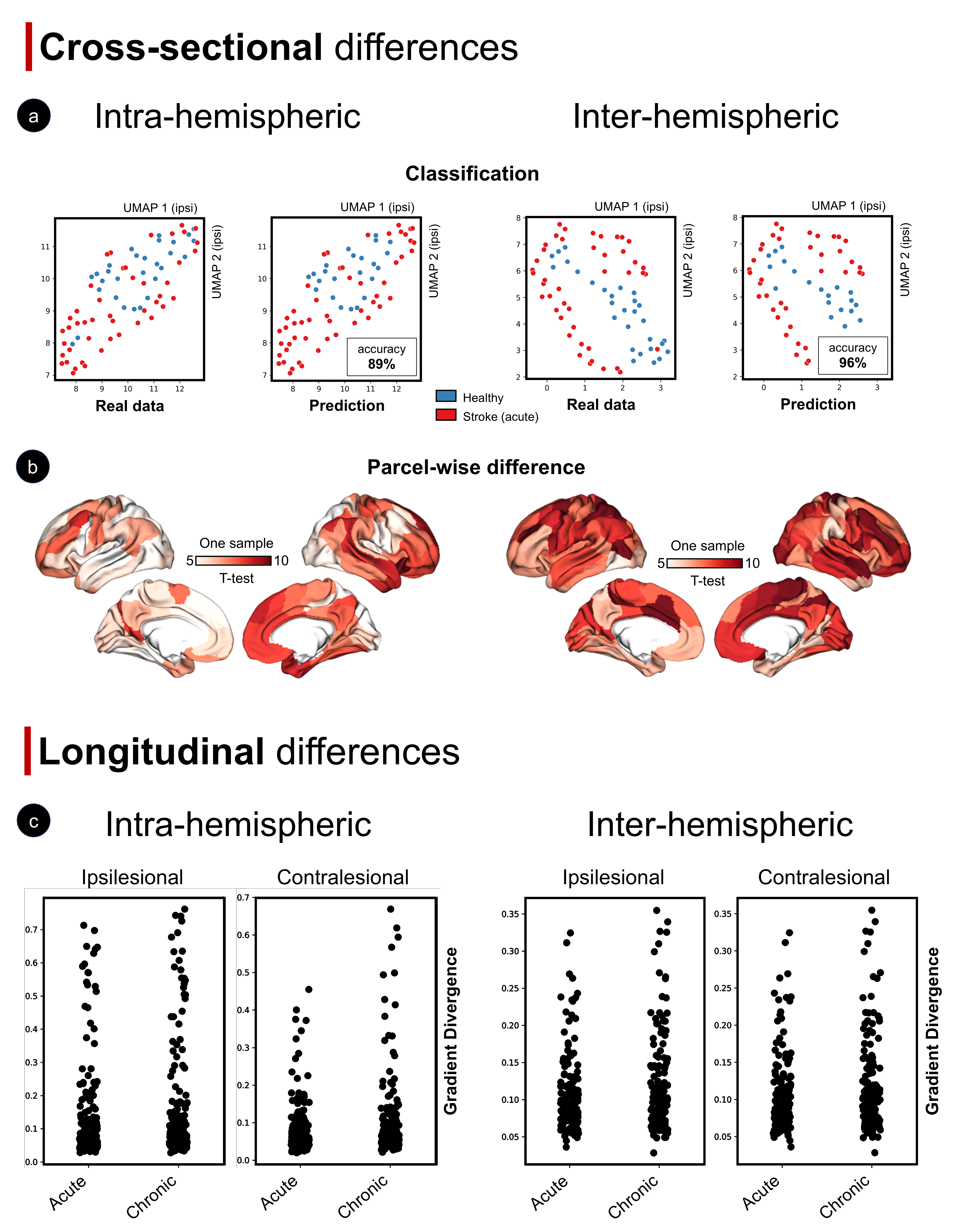

### Figure 6

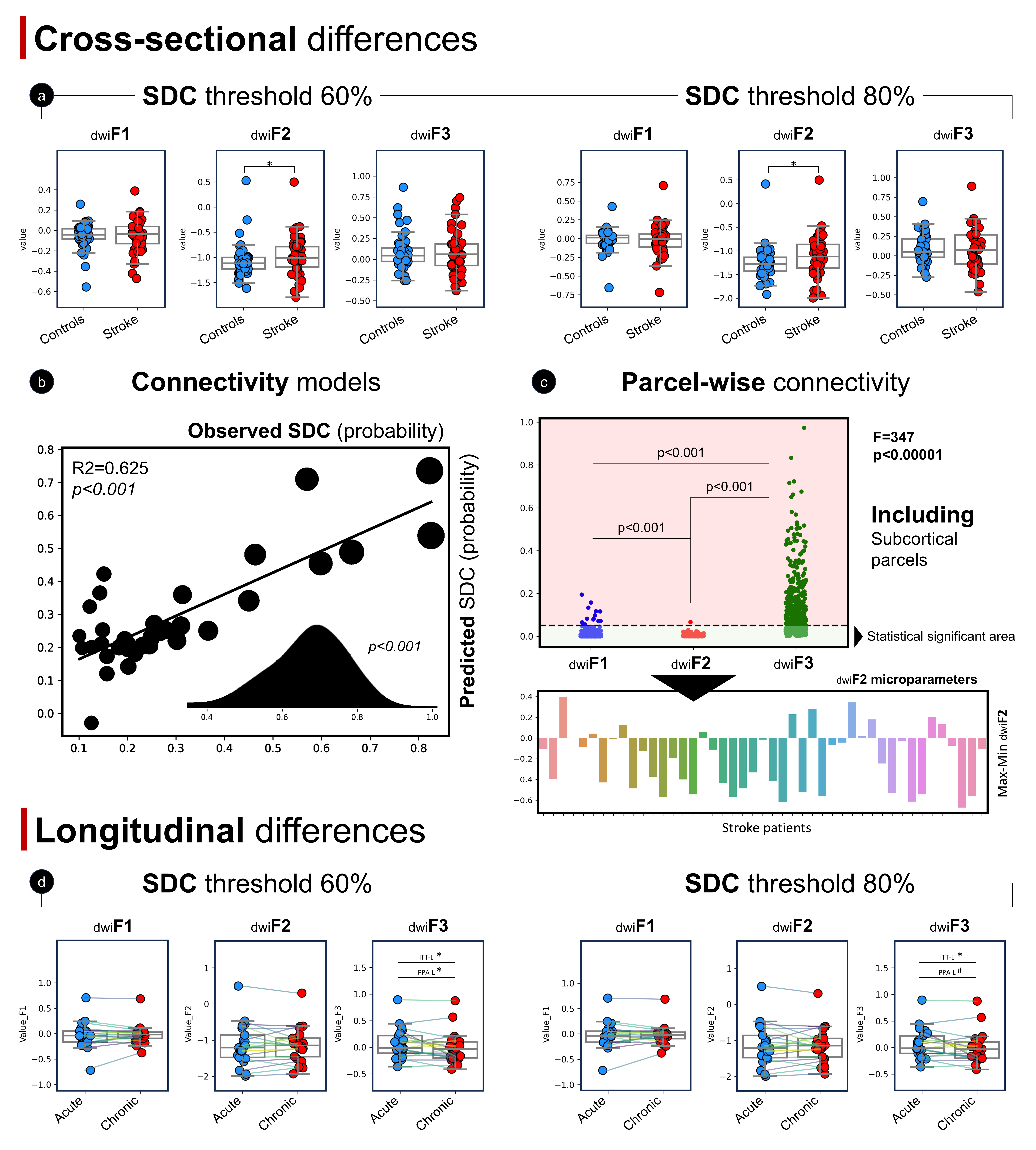

### Figure 7

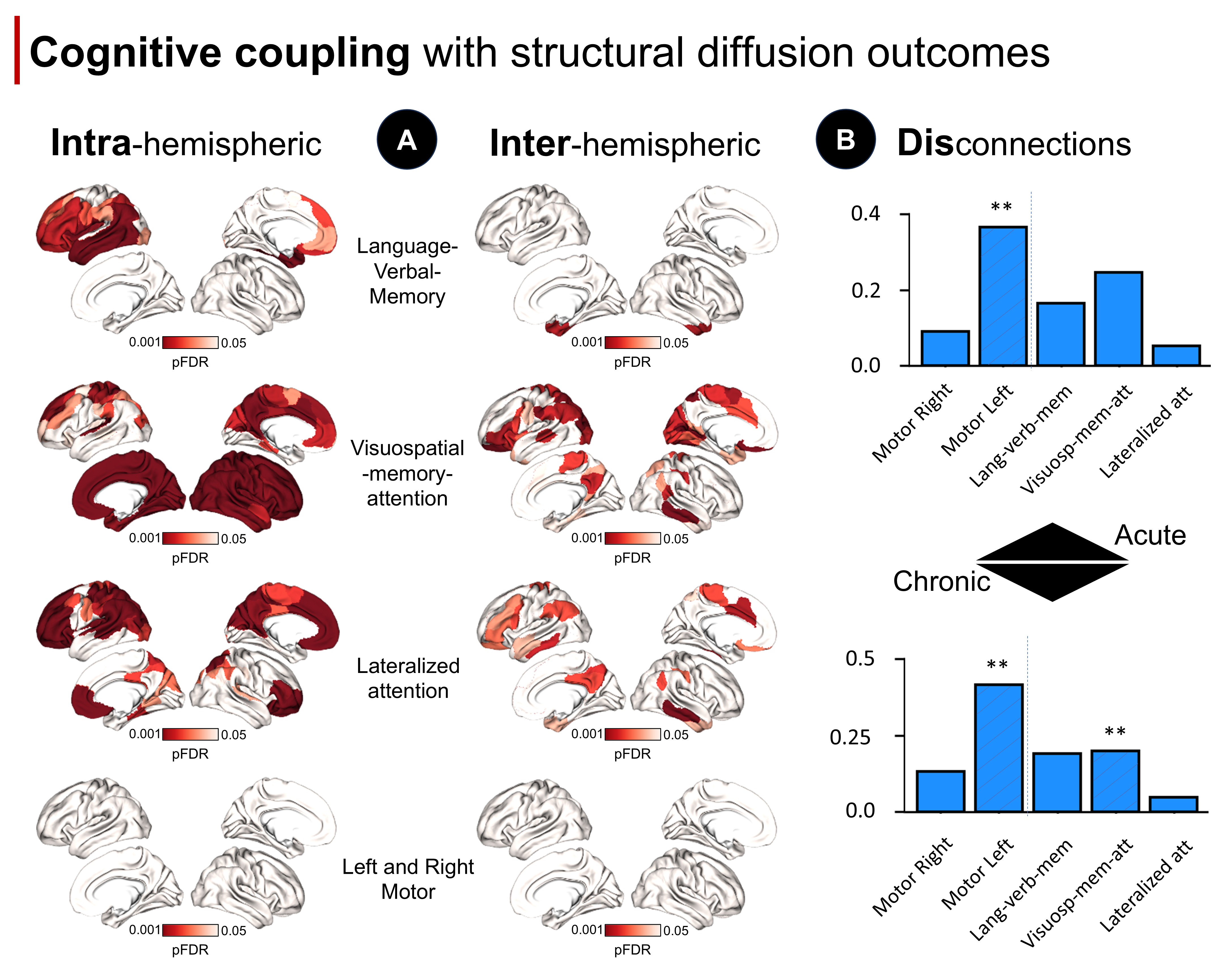
